# Comparative analysis of RNA expression in a single institution cohort of pediatric cancer patients

**DOI:** 10.1101/2024.09.25.24314005

**Authors:** Yvonne Vasquez, Holly Beale, Lauren Sanders, A. Geoffrey Lyle, Ellen T. Kephart, Katrina Learned, Drew Thompson, Jennifer Peralez, Amy Li, Min Huang, Kimberly Pyke-Grimm, Sofie Salama, David Haussler, Isabel Bjork, Sheri L. Spunt, Olena M. Vaske

**Author notes:** Contributed equally (Yvonne Vasquez, Holly Beale). Co-senior authors (Isabel Bjork, Sheri L. Spunt, Olena M. Vaske).

## Abstract

**Importance:** Genomic analyses solely focused on detecting mutations do not benefit most pediatric cancer patients. Alternate genomic approaches are needed to identify additional treatment biomarkers and therapeutic targets.

**Objective:** To evaluate the performance of our Comparative Analysis of RNA Expression (CARE) pipeline in nominating druggable targets in pediatric patients with difficult-to-treat or rare cancers.

**Design, Setting, and Participants:** Our cohort study, the Comparative Analysis of RNA Expression to Improve Pediatric and Young Adult Cancer Treatment (CARE IMPACT), was conducted collaboratively by the UC Santa Cruz Treehouse Childhood Cancer Initiative and Stanford University School of Medicine. From June 4, 2018, to September 24, 2020, UCSC Treehouse obtained and processed RNA sequencing (RNA-Seq) data for 35 tumor samples from 33 children and young adults with a relapsed, refractory or rare cancer. Each tumor RNA-Seq dataset underwent CARE analysis to reveal activated cancer driver pathways and nominate treatment options. We compare the CARE pipeline to other gene expression outlier detection approaches and discuss challenges and opportunities for the clinical implementation of RNA-Seq-based gene expression outlier detection for pediatric cancer patients.

**Exposures:** Patients underwent tumor RNA-Seq analysis and standard-of-care tumor DNA profiling. UCSC Treehouse compared each patient’s tumor RNA-Seq profile with over 11,000 uniformly analyzed tumor profiles from public data repositories. These comparisons reveal candidate cancer genes and pathways that represent potential therapeutic targets.

**Main Outcome(s) and Measure(s):** Proportion of patients for whom CARE provided useful clinical and biological information for patient care. Impact of comparator cohort choice on outlier findings.

**Results:** Of our 33 patients, 31 (94%) had CARE IMPACT findings of potential clinical significance. These findings were implemented in 5 patients, 3 of which had defined clinical benefit. We demonstrated that composition of comparator cohorts determines which outliers are detected and that large and diverse cohorts containing data from tumors similar to the patients produce the most clinically relevant outlier results.

**Conclusions and Relevance:** Comparative RNA-Seq analysis may identify additional cancer driver pathways and druggable targets in patients with rare or difficult-to-treat pediatric cancers relative to standard-of-care DNA profiling. This study highlights the clinical utility of CARE for pediatric tumors and underscores the need for further evaluation of this approach to improve patient outcomes.

**Key Points:** *Questions:* Is tumor RNA expression information useful for the clinical care of children with difficult-to-treat or rare cancers? How does choice of comparator cohort impact RNA expression results?

*Findings:* In this cohort of 33 pediatric patients, Comparative Analysis of RNA Expression provided useful clinical information for all patients; three of five patients who received treatment derived clinical benefit. We demonstrate the impact of comparator cohort composition on RNA outlier analysis.

*Meaning:* Tumor RNA expression information reveals useful information for the clinical care of pediatric cancer patients. Choice of comparator cohort size and composition impacts gene expression outlier detection.

## Introduction

Although overall clinical outcomes for pediatric cancer patients have improved over the past few decades, patients with recurrent/refractory or rare tumors still fare poorly.^1^ DNA-mutation-guided therapies have improved outcomes for some pediatric cancers.^2^ However, mutation analysis alone is often insufficient to identify therapeutic targets in most pediatric cancers because of the low incidence of clinically actionable mutations.^3,4^ This emphasizes the need for alternate genomic approaches to identify additional treatment biomarkers and therapeutic targets.

Several studies have demonstrated the utility of RNA sequencing (RNA-Seq) combined with DNA mutation analysis for pediatric cancer patients, primarily to identify clinically actionable variants^5,6^ or fusion transcripts.^2,7–13^ Studies in adult cancers^14^ have begun to look at abnormal gene expression to identify overexpressed pathways and targets for treatment. However, this approach is underexplored in pediatric cancers because the interpretation of gene expression results requires either matched normal tissues or comparator cohorts, which are harder to obtain for pediatric tumors.^15^ Additionally, interpretation and integration of genomic data into clinical care, requires a close partnership of multiple professionals, which can be challenging due to funding constraints, regulatory barriers and limited interoperability of medical systems.

Several multi-tumor-type pediatric cancer precision medicine studies^6,16–18^ have utilized RNA-Seq-derived gene expression to identify druggable genes and pathways that are highly expressed in patient tumor samples (termed gene expression outliers). However, they utilized inconsistent methods to identify such outliers, leading to difficulties in comparing across studies. The main inconsistency in identifying gene expression outliers is the choice of comparator cohort to assess abnormal gene expression in a patient’s tumor sample. For instance, Zero Childhood Cancer^17^ and INFORM^16^ utilized all other patients in the study as comparator cohorts to define gene expression outliers, while the Personalized Onco-Genomics (POG)^18^ Program utilized The Cancer Genome Atlas (TCGA) tumor and normal datasets.^19^

We previously described Comparative Analysis of RNA Expression (CARE), a comparative RNA-Seq approach for identifying overexpressed genes and pathways in pediatric tumors.^20^ This approach relies on large shared genomic datasets consistently processed for combined analysis. Unlike most other implementations of outlier detection, CARE compares expression in the focus sample to multiple cancer cohorts. In case studies, we have demonstrated the clinical utility of the CARE approach for both identifying treatments^21,22^ and for refining diagnoses of rare tumors.^23^ Here, we evaluate our approach in a cohort of 33 pediatric, adolescent, and young adult patients (age at diagnosis < 30 years) with recurrent/refractory or rare cancer treated at a single institution (Appendix Table A1). We demonstrate that comparative RNA-Seq analysis was informative for most study participants, including three of the five patients who received the identified therapy and derived clinical benefit. We explore the impact of comparator cohort composition on gene expression outlier analysis and highlight the importance of both comparator cohort size and composition. This study underscores the added value of gene expression profiling in pediatric oncology and highlights their unique challenges.

## Methods

### Study design and patients

The Comparative Analysis of RNA Expression to Improve Pediatric and Young Adult Cancer Treatment (CARE IMPACT) study (Figure 1A) was conducted collaboratively by the UC Santa Cruz (UCSC) Treehouse Childhood Cancer Initiative and Stanford University School of Medicine. Patients under 30 years of age with a known or suspected recurrent/refractory solid tumor or relapsed leukemia undergoing tumor sampling as part of their standard care were eligible. Patients with a newly diagnosed high-risk cancer for which there was no established standard of care were also included.

**Figure 1.**
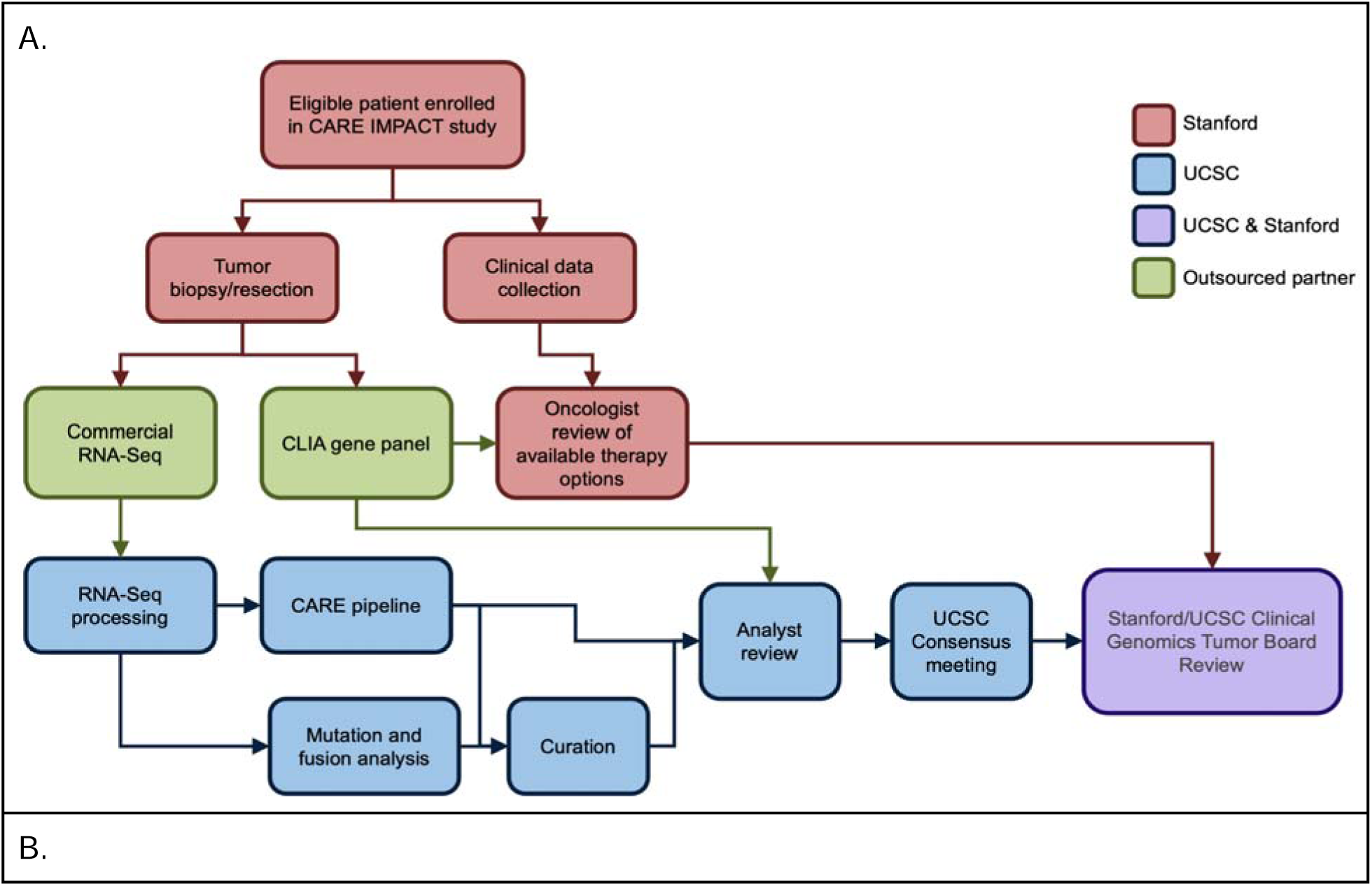
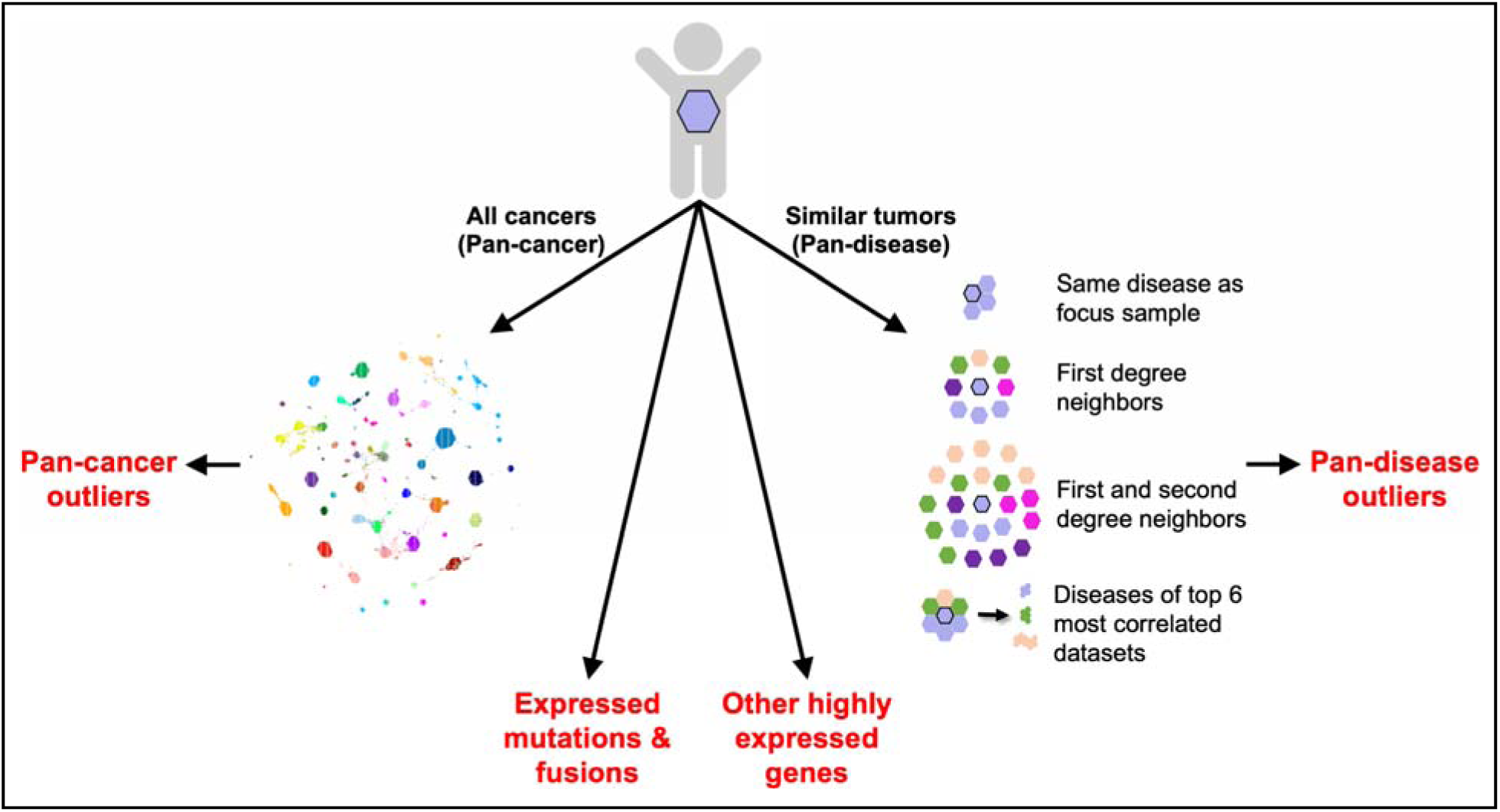
CARE IMPACT study workflow. **A**. The CLIA gene panel ordered was either a Foundation Medicine gene panel (https://www.foundationmedicine.com/portfolio) informed by the patient’s diagnosis or a Stanford Solid Tumor Actionable Mutations Panel (STAMP)(https://stanfordlab.com/content/stanfordlab/en/molecular-pathology/molecular-genetic-pathology.html/). All study components are described later in the manuscript or the supplemental methods. **B.** CARE IMPACT pipelines for identifying tumor vulnerabilities. CARE identifies gene expression outliers in each patient’s tumor (hexagon) relative to all other tumors in a large compendium (pan-cancer analysis) and to a subset of the compendium restricted to tumors with similar RNA expression and/or histology (pan-disease analysis). For pan-disease analysis, the focus sample is compared to four cohorts to identify outliers. If an outlier is detected by at least two pan-disease cohorts, it is considered a consensus outlier. Fusion and RNA variant pipelines are also applied to identify expressed mutations and fusions.

Prior to any study procedures, informed consent (from patients over 18 years of age or the patient’s legal guardian for those under 18 years of age) and assent (from patients 7 to 18 years of age) were obtained according to institutional guidelines. Before initiation, this study was approved by both the Stanford and UCSC Institutional Review Boards. UCSC Treehouse researchers never received direct patient identifiers during the duration of this study. Instead, de-identified clinical data was sent to UCSC investigators and secondary Treehouse identifiers (TH34_XXXX_S0X) were generated that could not be linked to direct patient identifiers. De-identified clinical data retrieved from each patient’s medical record included age, sex, race, ethnicity, cancer diagnosis, disease features, and treatment history.

Each patient’s tumor underwent standard-of-care DNA mutation analysis and CARE analysis (Supplemental Methods, Figure 1B).

### Clinical genomics tumor board meetings

CARE IMPACT findings were discussed in interdisciplinary, interinstitutional clinical genomics tumor board meetings where the treating oncologist presented the patient’s medical history, goals of care, and therapies being considered. Discussion focused on the strength of the analytical findings, the clinical evidence available to support the use of each identified treatment, and how to prioritize each option in the context of other available treatment options (Supplementary Methods). All findings were divided into “Accepted” and “Declined” based on how clinically useful the clinical team perceived them to be (Supplemental Methods). For patients who received a treatment nominated by the CARE IMPACT analysis, therapeutic benefit was defined as stable or decreasing evidence of disease >6 weeks after initiation of the treatment based on the treating oncologist’s assessment of relevant clinical, pathologic, and imaging studies. Patients with therapeutic benefit were followed for disease and survival outcomes.

### Assessment of comparator cohort impact on outlier detection

To determine whether the choice of comparator cohort influenced the outlier analysis in a clinically meaningful way and to compare gene expression outlier approaches used in other precision medicine studies incorporating RNA-Seq,^16–18^ we assessed the results of outlier detection with four different pre-defined comparison cohorts: the full compendium (equivalent to CARE pan-cancer analysis, n=12,747), all TCGA datasets (n=9806), data from pediatric patients (age at diagnosis < 30 years, n=2814), and data from all cases from a single institution cohort (Stanford, n=110). Of the 12,747 RNA-Seq datasets in the compendium, 9806 (76.9%) are from TCGA; of those, 9440 (96.3%) are adults. Of the 2941 non-TCGA datasets, 96.8% are pediatric (age at diagnosis < 30 years). This analysis was based on automated findings using the most recent compendium and CARE IMPACT version; no curation was performed.

## Results

### Patient characteristics

Thirty-three eligible patients were enrolled in CARE IMPACT between March 2018 and August 2020 (Appendix Table A1); 32 patients had a recurrent/refractory tumor, and one had a newly diagnosed high-risk cancer without an established standard of care. The median age at diagnosis was 11 years (range 0-24 years); 55% were male. Soft-tissue sarcoma was the most frequent cancer subtype (n=16, 48%).

### CARE IMPACT findings

CARE IMPACT analysis of the 35 tumors identified 89 clinically relevant findings presented in clinical genomics tumor boards (Appendix Table A3). Of these findings, 32 (36%) were uniquely identified by the automated pan-cancer pipeline, 9 (10%) were uniquely identified by the automated pan-disease pipeline (canonical consensus), 8 (9%) findings were uniquely identified by the automated pan-disease pipeline after adding a curated pan-disease cohort (curated consensus), 11 (12%) findings were uniquely identified by other means involving curation (mutations, fusions, other highly expressed genes, single cohort pan-disease outliers), and 29 (33%) findings were identified by both pan-cancer and pan-disease pipelines (Figure 2A and Appendix Table A3).

**Figure 2.**
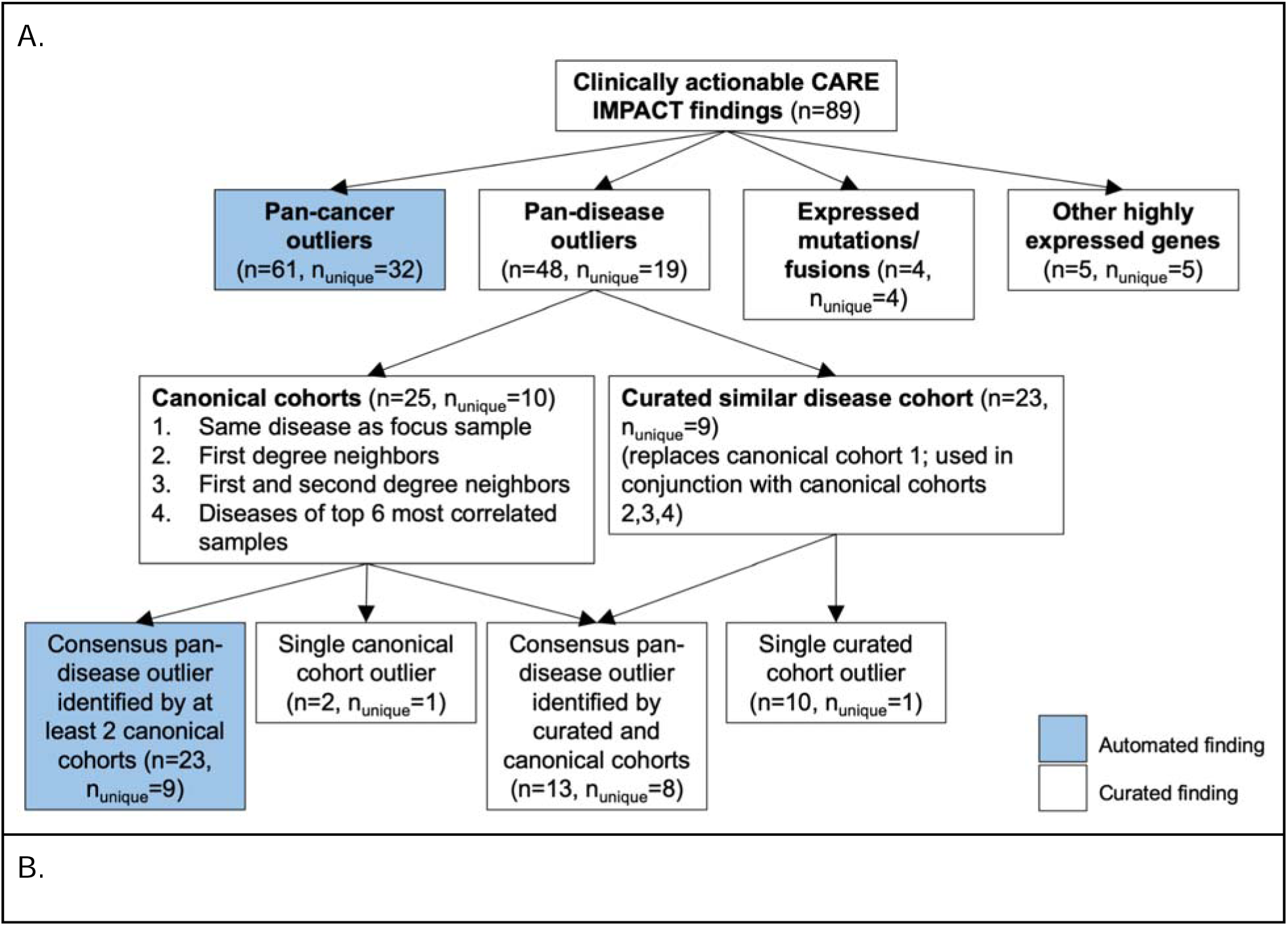
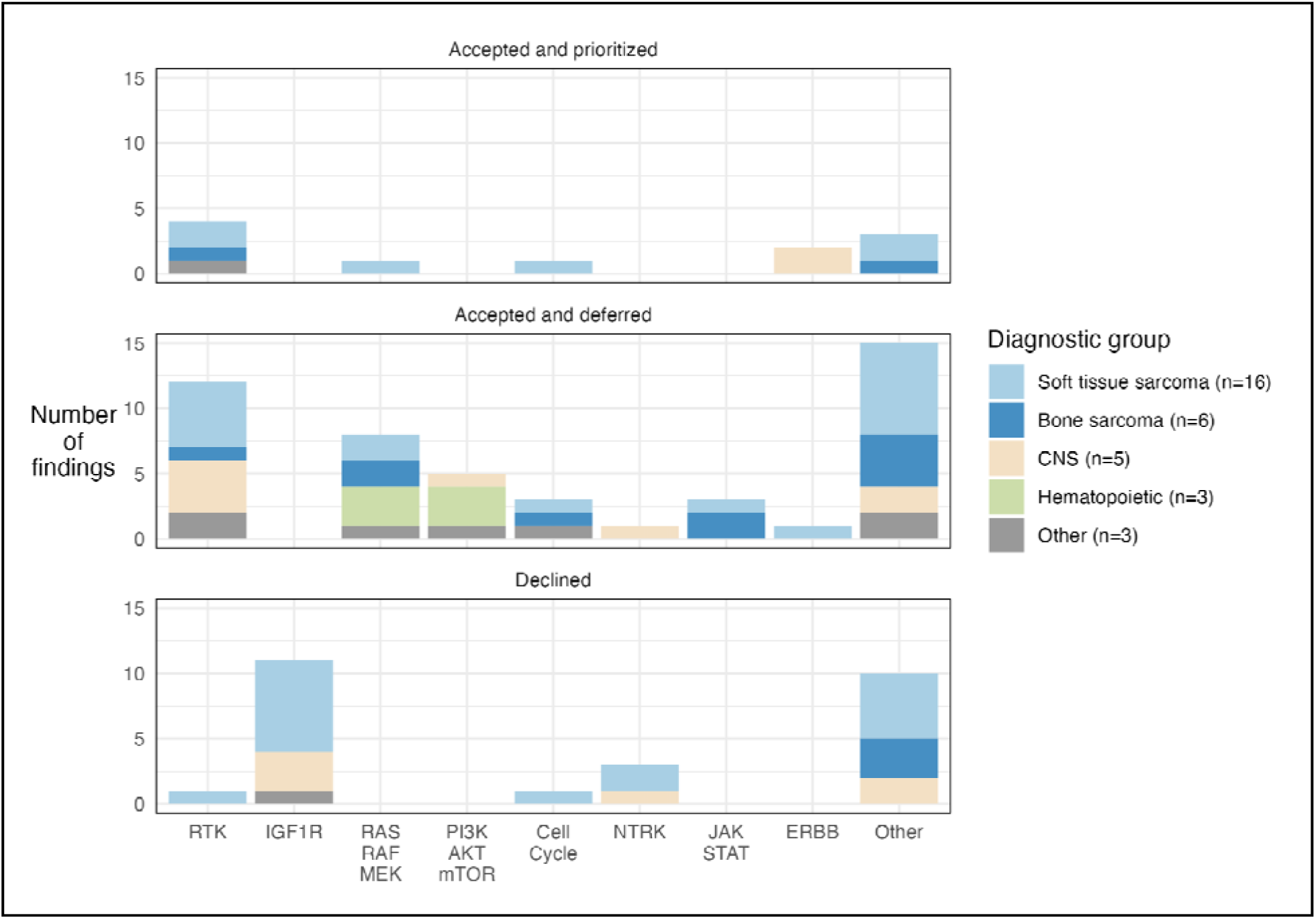
CARE IMPACT findings summarized. **A.** Breakdown of 89 clinically relevant CARE IMPACT findings by method of detection. The automated CARE IMPACT pipeline and human curation identified 89 findings with evidence from four possible sources: pan-cancer outliers, pan-disease outliers, expressed mutations/fusions, or other highly expressed genes. A fraction of findings were identified by both the pan-cancer and pan-disease analyses. The total number of RNA-Seq findings that fell into each category are enumerated in parentheses for each finding type. Findings that were uniquely identified by only the method listed in the box are designated by n_unique_. Pan-disease findings are further categorized by the cohorts used and the number of pan-disease cohorts used to detect them. Consensus outliers were defined as those identified by at least two cohorts. **B.** Breakdown of 89 CARE IMPACT findings by tumor vulnerability category and prioritization status for 33 patients studied. Tumor vulnerability category is defined as the category a gene falls under in terms of its function. Each bar graph represents a prioritization status designated by a clinician (Supplemental methods). The bars show total counts of CARE IMPACT findings in each tumor vulnerability category, colored by diagnostic group. The number of patients in each diagnostic group is indicated in the legend. Receptor Tyrosine Kinase (RTK).

Of the 89 CARE IMPACT findings identified and reported to Stanford, 70 (79%) were identified by our automated CARE pipelines, and 19 (21%) were identified only by human curation. Curation identified additional findings for 13 patients, three of whom had no findings identified by the automated pipeline.

All patients had at least one clinically relevant CARE IMPACT finding identified (Appendix Table A3, Figure 2B). While in over 10 cases, IGF1R was identified as a druggable gene expression outlier; it was not considered a useful finding because of the broad failure of IGF1R inhibitors in clinical trials.^24^ In contrast, gene expression outliers in the other vulnerability categories, notably Receptor Tyrosine Kinases (RTKs), were considered useful because of the availability of clinical data on the inhibitors.

### CARE IMPACT treatment outcomes

For CARE IMPACT findings nominated for each patient, we assessed the clinical relevance, how the patient’s care was affected, and response to therapy for those who received treatment (Appendix Figure A1, Appendix Table A3). CARE IMPACT findings provided helpful information for selecting treatment in five cases (Table 1, Supplemental Methods). Three patients had a response of stable disease after two months of treatment that was maintained for at least six months, and two had disease progression (Table 1, Figure A1). One of these patients was ultimately rendered disease-free with an identified therapy and surgery, as described in a separate manuscript.

**Table 1.** Patients treated based on CARE IMPACT findings. Five patients were treated based on CARE IMPACT findings. The response to therapy and the status of the patients after treatment are described. (NED -no evidence of disease, AWD - alive with disease, DOD - dead of disease, SD - stable disease, PD - progressive disease)

| Patient ID | Disease | Gene target | Finding type | Pan-disease type | Cohort type | Drug given | Therapy response (duration) | Current status after treatment discontinuation (duration) |
| --- | --- | --- | --- | --- | --- | --- | --- | --- |
| TH34_1 349 | gastrointestinal stromal tumor | KIT | pan-cancer, pan-disease | single cohort | curated | sunitinib | SD (37 mo); therapy discontinued due to insurance issues. PD | AWD (1.9 yrs) |
| TH34_1 352 | myoepithelial carcinoma | FGFR1, FGFR2, PDGFR A | pan-cancer, pan-disease | single cohort | canonical | pazopanib | PD | NED (2.2 yrs) |
|  |  | CCND2 | pan-disease | single cohort | canonical | ribociclib | SD (12 mo), followed by complete resection of residual tumor and 12 additional months of the same therapy |  |
| TH34_1 381 | ependymoma | ERBB2 | pan-disease | consensus | canonical | neratinib | PD | DOD |
| TH34_1 456 | osteosarcoma | KDR/VEGFR2 | pan-cancer, pan-disease | consensus | canonical | cabozantinib | SD (11 mo), then PD | DOD |
| TH34_2 351 | embryonal rhabdomyosarcoma | MAP2K1 | 95th percentile |  |  | trametinib | PD | DOD |

In five additional cases, the clinician was interested in administering a treatment supported by the CARE IMPACT analysis, but it was ultimately not used due to rapid disease progression (n=3), unavailability of the therapy (n=1), or because the family requested therapy initiation before an investigational new drug (IND) application could be completed (n=1) (Figure A1, Supplemental Methods).

In 21 cases, the CARE IMPACT findings deemed as most helpful information by the clinician were deferred because the treating oncologist elected a treatment option with more published evidence of efficacy (Figure A1). In 16 cases, the primary reason for deferring the identified treatment was the existence of another treatment with more published data in the specific condition (Supplemental Results). In three cases, the CARE IMPACT findings were deferred because the patient had not yet received the known standard of care treatment for their disease. In these cases, the clinician indicated they would consider using the identified treatment if the standard therapy was ineffective. The patient no longer needed treatment in two cases.

For two patients, the clinician did not find any of the nominated CARE IMPACT findings informative for treatment. In one case, the clinician was aware of studies in the cancer being treated showing limited efficacy of the drug identified in the CARE IMPACT analysis. In another case, all findings lacked FDA-approved.

Human curation identified informative findings for which the patient received therapy in three cases. Of those three patients, one achieved stable disease, one achieved no evidence of disease, and one had progressive disease on the CARE IMPACT elected therapy (Table 1, “single cohort” pan-disease type).

### Comparison of outliers detected by alternate comparator cohorts vs CARE analysis

While other pediatric precision medicine studies^6^ have utilized gene expression outlier analysis to identify targets for therapy, there is no consistency in how the outliers are defined in terms of the composition of comparator cohorts. To evaluate the impact of cohort composition on the outlier results, we compared the outliers detected by CARE to outliers detected using other common outlier detection strategies (e.g. single study cohort,^16^ TCGA cohort,^18^ a diversity of pediatric and young adult tumors^17^).

The automated CARE analysis identified 89 clinically relevant outliers (Figure A2, Table A10; summarized in Table 1 below). Seventy-two percent of the outliers had pathway support (Table 2, Supplemental Methods). Outliers were detected in 33 of the 35 analyzed tumor datasets and in 31 of the 33 patients. Of the three alternative cohorts used for outlier detection, comparisons to the TCGA cohort best replicated the automated CARE results, identifying 82% of the outliers, followed by a single institution cohort (Stanford) (43%) and the pediatric cohort (22%). Most of the automated CARE outliers detected by comparison to the Stanford cohort or the pediatric cohort were also detected by comparison to TCGA (Figure A3).

**Table 2.** Pathway support status of outliers detected by different comparative cohorts.

| **Comparison cohort** | **Number of CARE druggable outliers detected** | **Number of CARE druggable outliers with pathway support** | **Fraction of CARE druggable outliers with pathway support** |
| --- | --- | --- | --- |
| Treehouse CARE total | 89 | 64 | 64/89 (72%) |
| Treehouse CARE pan-cancer only | 72 | 47 | 47/72 (65%) |
| Treehouse CARE pan-disease only | 38 | 29 | 29/38 (76%) |
| Stanford | 38 | 25 | 25/38 (66%) |
| TCGA | 73 | 53 | 53/73 (73%) |
| Pediatric | 20 | 10 | 10/20 (50%) |

### TCGA-only cohort is of limited use for outlier analysis of pediatric samples

We next considered outliers detected solely by approaches other than the automated CARE analysis (Figure 3A). Twenty-five of these outliers were detected using outlier analysis against the TCGA cohort, and most outliers (21/25, 84%) were uniquely detected in this comparison and not present in comparisons against the Stanford or pediatric cohorts. Of the 16 genes in which these 21 outliers were detected, all 16 have wider distributions of expression in pediatric cohorts compared to TCGA, leading to higher outlier thresholds in cohorts with pediatric datasets. For example, the *FGFR3* IQR is 3.69 log2(TPM+1) in the pediatric cohort and 3.05 in the TCGA cohort (Figure 3B). Even though the median *FGFR3* expression value is lower in the pediatric cohort than in the TCGA cohort (Table A11), the outlier threshold is higher due to the larger IQR. If we had used the TCGA cohort instead of the Treehouse compendia as the comparison cohorts in the CARE IMPACT study, we would have identified 98 gene expression outliers, 21 (21%) of which would not be relevant to pediatric cancers, as their outlier status would not be replicated if pediatric datasets were added to the comparator cohorts (Figure 3B).

**Figure 3.**
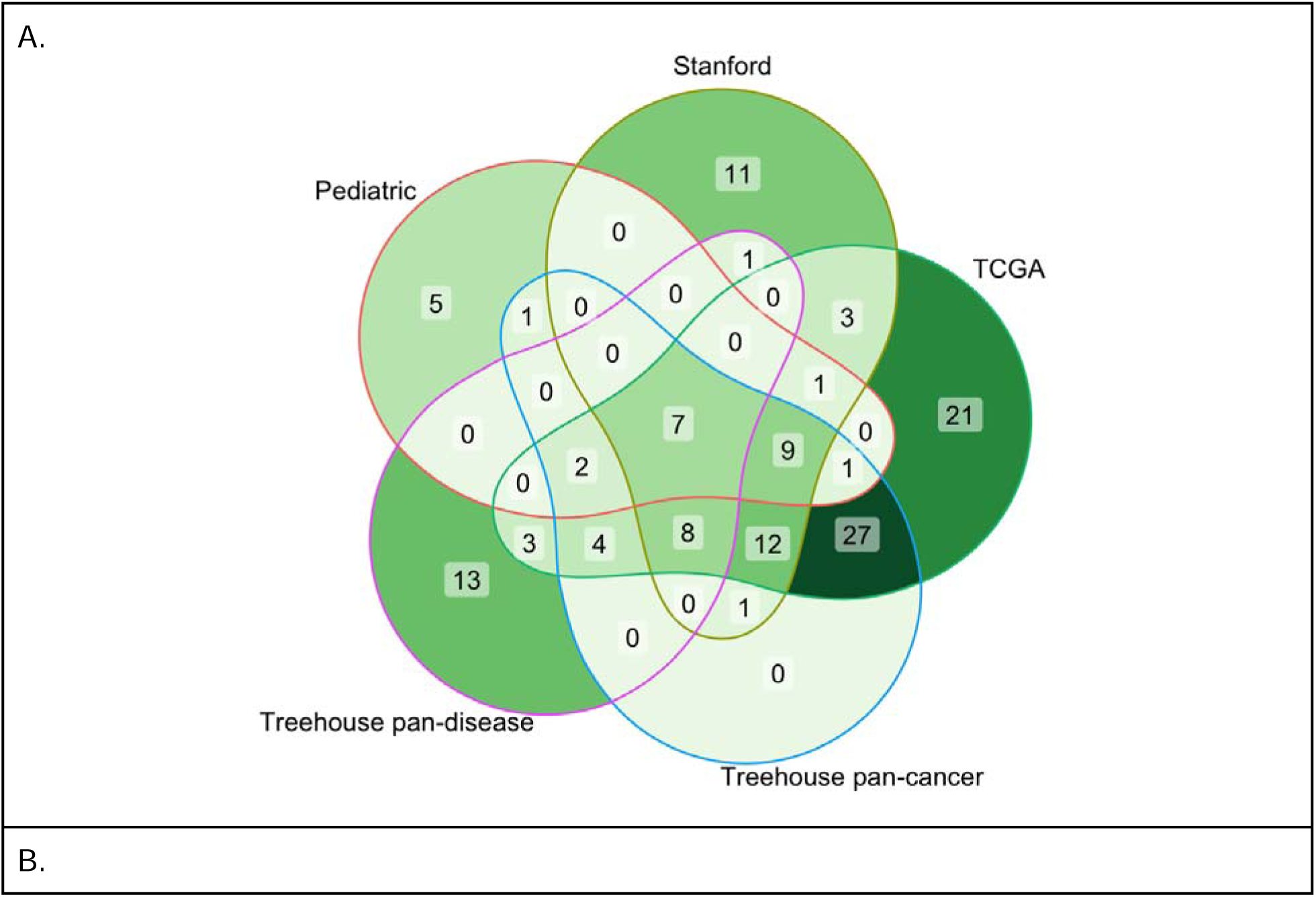
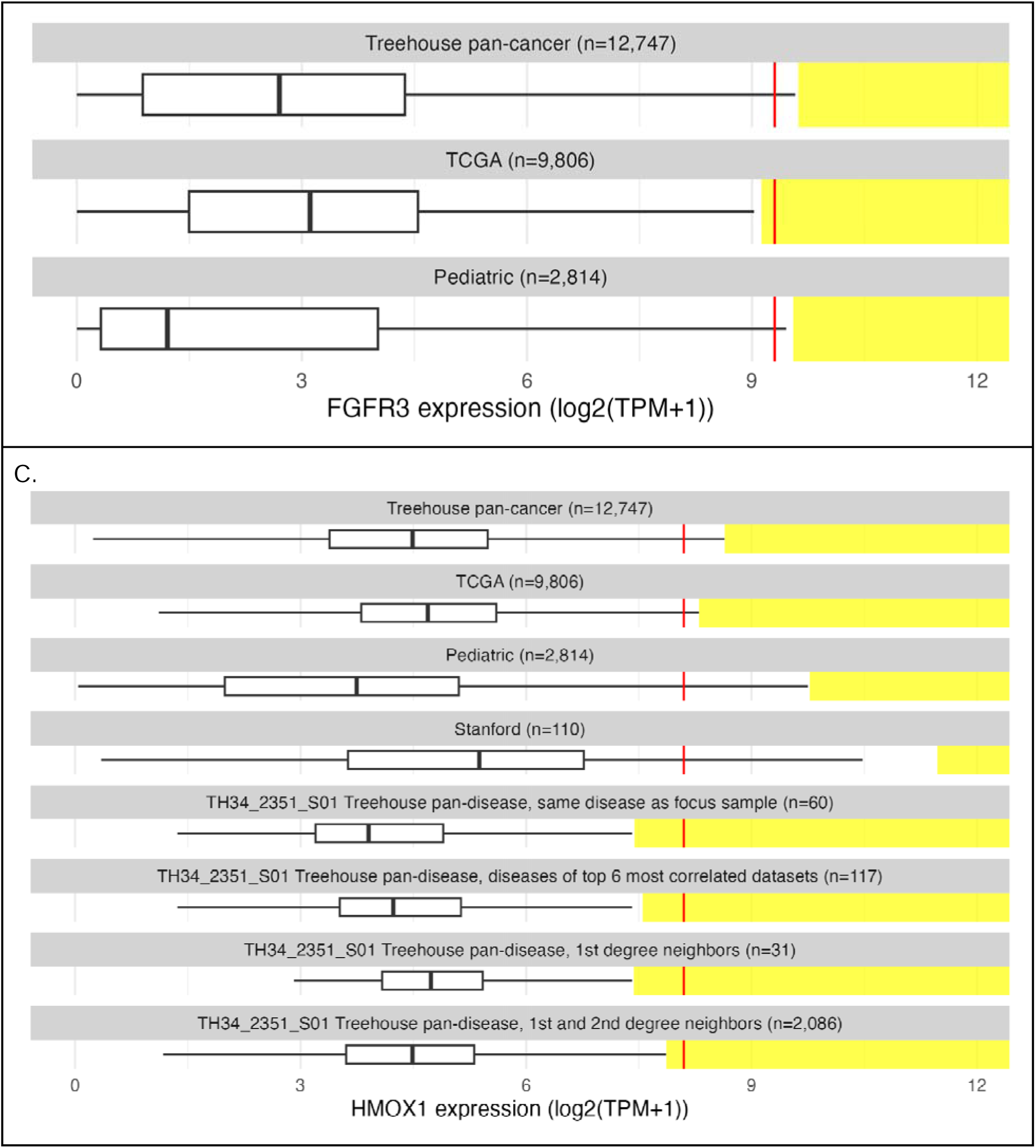
The choice of comparator cohort affects the outlier status. **A.** Numbers of outliers detected relative to the comparator cohort(s) used. The total number of outliers detected relative to each combination of cohorts is displayed in each intersecting region. The largest set consists of 27 outliers detected relative to both the Treehouse pan-cancer cohort and TCGA. **B.** FGFR3 expression in the patient sample TH34_1455_S01 illustrates the impact of cohort selection on outlier status. The FGFR3 expression level in the sample is denoted with a vertical red line plotted with respect to the distribution of FGFR3 gene expression in log2(TPM+1) across the comparator cohort (x-axis). The outlier range is denoted with a yellow bar. **C.** HMOX1 expression level in the sample TH34_2351_S01 (red) relative to various comparator cohorts. Vertical red line denotes gene expression level in the sample with respect to the distribution of HMOX1 gene expression in log2(TPM+1) across the comparator cohort (x-axis). The outlier range is denoted with a yellow bar.

### Using sample-specific comparator cohorts increases findings by 11%

The CARE pan-disease analysis uses a personalized comparator cohort customized to the transcriptome of a patient’s tumor (Supplemental Methods). For example, *HMOX1* in TH34_2351_S01 was not identified as an outlier gene in any of the static cohorts (Treehouse compendium, TCGA, pediatric and Stanford cohorts; Figure 3C). However, *HMOX1* is expressed exceptionally highly compared to the personalized cohorts that are generated for this dataset: other embryonal rhabdomyosarcomas (“same disease”), transcriptionally correlated datasets (“First degree neighbors” and “First and second degree neighbors”; see Methods), and datasets with the same disease as one of the top 6 most correlated datasets. In total, Treehouse pan-disease analysis added 13 findings not detected by predefined cohorts, increasing the total number of findings by 11% from 117 to 130 (Figure 3C).

## Discussion

The mutational burden of childhood cancers is much lower than that of adult cancers, resulting in a lower frequency of molecular targets for therapy.^11^ In this cohort of 33 recurrent, refractory or rare pediatric tumors, implementation of RNA-Seq-based gene expression analysis in a clinical setting was feasible and produced informative molecular abnormalities in all patients. Of our 33 patients, 31 (94%) had CARE IMPACT findings of potential clinical significance. These findings were implemented in 5 patients, and in 3 out of 5, the treatments produced defined clinical benefit.

In addition to identifying novel druggable aberrations, comparative RNA-Seq can clarify which patients may benefit from a biomarker-targeted therapy in the absence of the established biomarker. For example, our patient with GIST (TH34_1349) and wild-type *KIT* had *KIT* overexpression and benefited from sunitinib (Supplemental Methods). Even though sunitinib is a known treatment strategy for wild-type GIST; however, not all wild-type patients benefit from the therapy.^25^ Therefore, KIT overexpression may serve as a biomarker of response to KIT inhibitors in the setting of wild-type *KIT* where KIT-targeted therapy might not otherwise be prioritized.

Despite most patients harboring an informative finding, the findings were only implemented in five cases, and often the suggested treatments were deferred in favor of other therapies. This highlights the challenge of evaluating the impact of genomics-guided treatments in populations with multiple treatment options and clinical trials available. For 19 of 33 (58%) of the patients with CARE IMPACT findings, other treatments with more data in the disease (including standard of care) were available for consideration by the clinical teams. Proving the value of genomics-guided treatments will depend on doing studies where patients receive the identified therapy.

Most tumors were analyzed after the standard of care treatments had been exhausted, leaving the patients prone to rapid clinical decline. For three patients in which RNA-Seq analysis identified a treatment that would have been implemented, the patients had rapid disease progression and died before they could receive the treatment. This emphasizes the need for timely integration of molecular analysis in cancer care.

This study is limited by a relatively small cohort of heterogeneous pediatric diseases. Further examinations of clinical utility of comparative RNA-Seq in larger cohorts of single diseases are warranted. Furthermore, this study is limited by the lack of a clinically approved RNA-Seq assay. Clinical validation of this comparative RNA-Seq protocol, which is underway, will aid in further evaluation of the clinical utility of this approach in patient care.

A key challenge in the clinical implementation of RNA-Seq-based gene expression is standardizing gene expression outlier analysis. We demonstrated that the composition of comparator cohorts determines which outliers are detected and that large and diverse cohorts containing data from tumors similar to the patient’s produce the most clinically relevant outlier results. Comparing pediatric datasets to TCGA-only cohorts produces gene expression outliers with limited relevance to pediatric cancers, i.e. the identification of gene expression that is exceptionally high for adult cancers but not for pediatric cancers. Approximately one-fifth of the outliers detected when comparing to TCGA only (which is >96% adults) are not detected in comparison to the other cohorts, which contain a minimum of 22% pediatric datasets, indicating that these outliers are due to the paucity of pediatric samples in the TCGA cohorts.

In addition, we show that our pan-disease analysis, which compares a dataset to dynamically generated, patient-specific cohorts based on disease and molecular similarities, generates orthogonal results. Of the 38 pan-disease findings, 34.2% were not detected by any predefined single-cohort analysis. Pan-disease findings identify how the patient’s tumor differs from other similar tumors, which may highlight potential therapeutic alternatives for patients whose disease does not respond to the standard-of-care treatment. Therefore, an ideal comparator cohort would be composed of hundreds of datasets of each pediatric and adult tumor type. Important limitations to constructing large comparator cohorts for gene expression outlier analysis are the siloing of RNA-Seq data and the differences in the processing and analysis of RNA-Seq datasets, hindering the merging of multiple datasets.^26,27^ We anticipate that NCI’s Childhood Cancer Data Initiative (CCDI) will help solve the data siloing dilemma by creating a federated framework in which RNA-Seq data could be shared across stakeholders.

The incorporation of RNA-Seq-based expression analysis to identify clinically relevant therapeutic targets in difficult-to-treat pediatric tumors is feasible as a collaborative effort of an interdisciplinary team. This approach revealed druggable aberrations in most of our cohort and can be performed within the time frame required for patient care. In all cases, we convened an interdisciplinary, interactive genomic tumor board tailored to a specific patient’s needs. This tumor board was highly educational to both clinicians and researchers and led to improvements in the analysis and reporting process (discussed in a separate manuscript). Therefore, we believe that close partnerships of multiple professionals are essential to a successful precision medicine program.

## Data Availability

All data produced are available online at https://treehousegenomics.ucsc.edu/public-data/

## Acknowledgments

This study was funded by the California Initiative to Advance Precision Medicine (CIAPM), Stanford Medicine Children’s Health and Stanford University School of Medicine, American Association for Cancer Research NextGen Grant for Transformative Cancer Research, Emily Beazley Kures for Kids Fund St Baldrick’s Consortium Grant, Unravel Pediatric Cancer, Team G Childhood Cancer Foundation, and Live for Others Foundation. Dr Haussler is a Howard Hughes Medical Institute Investigator. Dr. Vaske holds the Colligan Presidential Chair in Pediatric Genomics. Dr. Spunt held the Endowed Chair of Pediatric Cancer during the conduct of this study. We are grateful to Drs. Alejandro Sweet-Cordero, Avanthi Tayi Shah, Arun Rangaswami, Norman Lacayo, and Julien Sage for their contributions to the study. We thank the Stanford pediatric oncologists and other clinical staff for participating in clinical genomics tumor boards and other study activities. We are indebted to all the patients and families for their participation in the study.

