## Supplemental Appendix for "Comparative analysis of RNA expression in a single institution cohort of pediatric cancer patients"

**Supplementary Appendix**

### **Supplemental Methods**

#### **RNA sequencing protocol**

For each patient, fresh-frozen biopsy or resection samples taken at the time of enrollment or from earlier procedures were sent to Covance by Labcorp (Covance) for RNA sequencing (RNA-Seq). RNA was extracted with the Qiagen RNEasy kit. A sequencing library was prepared with the Illumina TruSeq Stranded mRNA Library Preparation and sequenced on an Illumina HiSeq 2500 sequencer to obtain 40-50 million reads.

#### **Patient data transfer**

De-identified clinical (Appendix Table A1) and mutation information (Appendix Table A4) were extracted from the medical record of each patient at study entry and sent to the UC Santa Cruz (UCSC) Treehouse Childhood Cancer Initiative for the analysis. De-identified raw RNA-Seq data files for Stanford patients were obtained by UCSC from Covance. Covance uploaded patient fastq files to UCSC Treehouse's encrypted Amazon Web Services (AWS) bucket and provided quality metrics. The files were downloaded from AWS to UCSC Treehouse's encrypted servers. RNA-Seq sample data with associated clinical metadata were managed using REDCap^1^ electronic data capture tools hosted at Treehouse. Patient age at diagnosis is reported in age range quartiles: 0-5, 6-10, 11-15, 16-20, and 21-25 years of age.

#### **Sequencing data analysis and CARE IMPACT computation pipelines**

From June 4, 2018, to September 24, 2020, UCSC Treehouse obtained and processed RNA-Seq data for 40 tumor samples from 38 children and young adults. Six samples were not included in the study because they didn't pass QC checks described later.

The RNA-Seq analysis (<https://github.com/UCSC-Treehouse/pipelines>) was uniformly performed as described previously,^2^ with the following modifications. The most recent docker for the UCSC Treehouse RNA-Seq analysis pipeline was used (docker command: docker pull quay.io/ucsc_cgl/rnaseq-cgl-pipeline:3.3.4-1.12.3).^3^ For this study, the geneBody_coverage.py tool was not run.

As part of the CARE IMPACT analysis, the automated CARE pipeline was employed (<https://github.com/UCSC-Treehouse/CARE>) to identify clinically relevant oncogenes and oncogenic pathways in each case. For the purposes of this study, clinically relevant genes were designated as genes whose products could be directly or indirectly targeted through the downstream signaling pathway by an approved drug or an investigational agent in any phase of clinical development (Appendix Table A2). Publicly available Treehouse polyA compendia (https://treehousegenomics.soe.ucsc.edu/public-data/) were used for contextual analysis of each patient (Appendix Table A3). CARE compares an RNA-Seq profile from a focus sample to comparator cohorts selected from the Treehouse compendia and yields two outputs: (1) datasets molecularly similar to the focus sample and (2) genes that are abnormally expressed in the focus sample. Tumors are considered molecularly similar if the Spearman correlation between their expression profiles is above the 95th percentile of all pairwise correlations within the compendium. Abnormally expressed genes are those exceeding the outlier threshold for the comparator cohort. Outlier thresholds are defined using the Tukey outlier detection method ((Interquartile Range)(1.5) + 75% Quartile). For each focus sample, pan-cancer and personalized pan-disease outlier analyses are performed. Pan-cancer outliers are those exceeding the outlier threshold defined by the entire compendium at the time of the analysis (at least 11368 tumor RNA-Seq profiles from both adult and pediatric patients) (Appendix Table A3). Pan-disease outliers are genes with expression exceeding the outlier threshold from at least two of the four personalized pan-disease cohorts: 1) datasets from tumors with the same diagnosis as the focus sample, 2) molecularly similar RNA-Seq datasets (first degree neighbors), 3) first and second degree neighbors (first degree neighbors plus RNA-Seq datasets molecularly similar to them), and 4) datasets from diseases present among the top 6 most correlated datasets (Figure 1B). Pan-cancer and pan-disease outliers were analyzed for enrichment of downstream pathways and signaling networks containing genes that could be targeted by available therapies.

In addition to the CARE pipeline, variant calling and fusion detection pipelines were run on all datasets. Together, these pipelines produced a list of clinically informative findings for each focus sample, including gene overexpression outliers, expressed mutations, expressed fusions, and other highly expressed genes.

#### **CARE IMPACT curation**

In addition to the findings identified by the automated pipelines, results were reviewed by human analysts in several steps. Firstly, if no comparator datasets from the same disease were available for the pan-disease analysis described above, a curated cohort of clinically similar tumors was constructed, in consultation with the pediatric oncologist, to replace the "same disease as focus sample" cohort used in the pan-disease analysis (Figure 1B). For example, for a patient (TH34_1415) diagnosed with undifferentiated sarcoma NOS, we combined multiple types of soft tissue sarcomas. Secondly, after CARE was run, the analyst would review highly expressed genes (95th percentile) that did not reach the threshold for automated reporting as gene expression outliers but might be relevant based on prior knowledge, literature searches, or other genomic information. In this study, findings categorized as “generated using human curation” are those identified using curated similar disease cohorts; those present in only one of four pan-disease cohorts; highly expressed non-outliers implicated by mutation; and mutations and fusion genes.

#### **Sample quality control metrics**

A previously described quality control (QC) framework^4^ was used to ensure sufficient quality of the RNA-Seq data for identifying overexpressed oncogenes and pathways. This method relies on counting MEND reads (Mapped to human genome, Exonic, and Non-Duplicate). Filtering the total pool of reads in an RNA-Seq sample for MEND reads results in a subpopulation of reads that reflect the integrity and quantity of RNA in the sample and indicate whether the data can be used for robust gene expression quantification. Of the 40 RNA-Seq samples obtained, 34 passed QC and were included in the study. Six samples failed QC. For two donors, the first datasets produced failed QC, but we were able to include them in the study because a subsequent sample passed QC. Three other donors were excluded from the study because no datasets, initial or subsequent, passed QC. One QC fail sample (TH34_2292_S01) was also included in the study after additional analysis to determine the validity of all the reported outlier genes, because it was the only sample available from that patient. For this sample, the number of measured genes was low and the majority of sequencing reads (95%) were duplicates. High numbers of duplicates can potentially cause log_2_(TPM+1) measurements to be inflated. Additional accuracy quantification analysis was run on this sample and outlier analysis repeated after removing all duplicates. For this QC fail sample, only oncogenes with expression greater than 8.5 log_2_(TPM+1) were selected for further analysis, to account for the high number of duplicate reads. A gene expression value of 8.5 log_2_(TPM+1) was chosen as the cutoff expression level because it was the 95th percentile of gene expression in the dataset.

#### **Datasets used for comparative RNA-Seq analysis**

Multiple versions of the Treehouse Gene Expression Reference polyA Compendium were used in this manuscript, each composed of RNA-Seq datasets derived from public repositories and our partner clinical sites (Appendix Table A5). Treehouse's gene expression compendia are publicly available (<https://treehousegenomics.soe.ucsc.edu/public-data/>).

#### **Gene lists used for pan-disease and pan-cancer analysis**

For the pan-disease analysis, 58581 genes from GENCODE Human Release 23 were used. For pan-cancer analysis an expression- and variance-filtered set of GENCODE 23 genes was used, as enumerated in Appendix Table A6. First, the expression filter drops any gene where 80% or more of the samples have an expression of 0. Second, the variance filter sorts the remaining non-dropped genes and sorts them by the variance of their expression level across the cohort. The 20% of these genes with the lowest variance are dropped regardless of absolute variance.

#### **Analysis of overexpressed genes**

Overexpressed gene lists for each patient sample were analyzed for enrichment of pathways and signaling networks containing genes that could be targeted by available therapies (Appendix Table A2).

We used the Drug Gene Interaction Database (DGIdb)^5^ to identify which overexpressed genes could be targeted by clinically available inhibitors. DGIdb is an open-source project that searches through publications and other curated databases for known or potential interactions between human genes and available inhibitors. To focus our findings on drug targets with known cancer relevance, we set DGIdb to query drug-gene interactions in the following four curated databases: CIViC, Cancer Commons, My Cancer Genome, and My Cancer Genome Clinical Trial. DGIdb does not contain all known drug-gene interactions, nor does it guarantee that any interaction is an appropriate therapeutic intervention. To address these limitations, we conducted additional literature review and consulted published clinical cancer genomic studies. We prioritized studies that considered gene expression information when assessing the druggability of each gene.

We used the Molecular Signature Database (MSigDB)^6^ to identify significantly overexpressed cancer pathways in each tumor sample by conducting gene set overlap analysis, which computes statistically significant pathways between the input gene list of overexpressed genes and the gene sets in the chosen MSigDB collections “Hallmark Gene Sets” and “Canonical Pathways”.

#### **RNA variant analysis**

This section of our data analysis pipeline (<https://github.com/UCSC-Treehouse/pipelines>) uses bam files generated from our RNA-Seq analysis pipeline for (1) alignment based variant detection and (2) variant annotation. Variants in a curated list of clinically relevant mutations (Appendix Table A7) are called using Freebayes^7^ (<https://github.com/freebayes/freebayes>) version v9.9.2-27-g5d5b8ac, by comparing the specific genomic loci in the reference and patient genomes. The list of variants outputted by Freebayes are annotated using SnpEff version SnpEff 4.3r.^8^ This information was used to complement available DNA mutation data. Of note, our list of clinically actionable mutations was updated once during the duration of this registry study. This pipeline has been dockerized and the code is available at <https://github.com/UCSC-Treehouse/mini-var-call>.

#### **RNA fusion analysis**

This pipeline uses a docker container generated by https://github.com/UCSC-Treehouse/fusion-for-core that runs STAR-Fusion^9^ on paired-end fastq files and filters the output against a list of known cancer fusion genes (Appendix Table A8). FusionInspector^10^ is run on the STAR-Fusion output for additional filtering and quantification. The filtering process requires that both fusion partners are in the known cancer fusion gene list. If there are no clinically relevant fusions in the filtered output, a data analyst reviews the unfiltered list for clinically relevant fusions involving promiscuous fusion partners.

#### **DNA mutation analysis and classification**

When adequate tumor tissue was available, a sample was sent for DNA mutation testing at either Foundation Medicine (https://www.foundationmedicine.com/portfolio) (FoundationOne Heme or FoundationOneCDx, as recommended for tumor type) or Stanford’s Solid Tumor Actionable Mutation Panel (STAMP)(https://stanfordlab.com/content/stanfordlab/en/molecular-pathology/molecular-genetic-pathology.html/) (Appendix Table A4). In one case (TH34_1447_S01), a tumor sample from a different metastatic site and time point in the patient’s cancer progression was sent for testing. Foundation Medicine and STAMP reports provide a list of variants classified as genomic (pathogenic) findings or as variants of uncertain significance (VUS). For reported pathogenic findings they are further classified as actionable if they have therapeutic implications. For actionable variants a table is provided listing potential therapies including FDA-approved therapies for patient’s tumor type, FDA-approved therapies in other tumor types, and potential clinical trials (Appendix Table A4). Tumor DNA mutation data was considered clinically useful for a patient if the mutation reports identified an actionable mutation that could be treated with an FDA-approved therapy, or a clinical trial was available at the time the report was generated. One caveat is that what mutation panels consider actionable changes over time with the rapidly developing field of cancer and clinical trials. For example, a NTRK1 variant was not reported as actionable by Foundation Medicine at the time of analysis, however, now could be considered actionable by TRK inhibitors. Of note, variants annotated by the testing site as "equivocal," meaning the amplification call is not definitive and should be confirmed by a second source, were still considered actionable if an FDA-approved therapy was listed. Additionally, in one case an activating KRAS mutation was listed as actionable with potential FDA-approved therapies listed that are known standard of care for the patient's disease type, however, the specific mutation was noted to render the patient resistant to therapy. We did not classify this variant as clinically actionable; however, it was still considered clinically useful because it could ultimately impact treatment decisions.

#### **Clinical genomics tumor board meetings**

Upon completion of the RNA-Seq data analysis, summary research reports were sent to the treating oncologist ahead of a Stanford registry study-specific clinical genomics tumor board meeting attended by the treating oncologist, additional pediatric oncologists (some of whom were part of the registry study team), genomics scientists, bioinformaticians, data analysts, nurse practitioners, a genetic counselor, and various trainees. This format allowed for rich interdisciplinary discussion of each case. Prior to each session, clinicians were asked to avoid using HIPAA-protected patient identifiers during case discussions to protect patient privacy. The treating physician presented the patient’s history, including past treatment, current medical status, goals of care, and potential therapies being considered. A Treehouse data analyst presented the RNA-Seq data, including specimen quality metrics, gene expression findings, targeted agents identified, and literature supporting or refuting the use of the targeted agent in the patient’s tumor or similar tumors. Available DNA mutation panel results were also presented. Discussion focused on both the analytical strength of the RNA-Seq findings (strength of overexpression, pathway support, supporting mutations) and the clinical evidence available to support the use of each targeted therapy, as well as how to prioritize each option in the context of other available treatment options. After discussion in the clinical genomics tumor board and any further analysis prompted by the discussion was complete, a final summary report was sent to the treating oncologist including molecular testing results, a TumorMap^11^ visualization of molecularly similar samples, clinically relevant overexpressed genes and pathways, and suggestions for targeted treatments.

#### **Assessment of clinical utility of CARE IMPACT findings**

At a timepoint >6 months from study enrollment, each patient's medical record was reviewed, and the treating oncologist was interviewed to determine the clinical utility of each CARE IMPACT finding. The clinical utility of each finding was categorized as follows and is displayed in Figure A1:

1) Accepted and prioritized - The targeted agent was FDA-approved, had published phase I safety data in children, and/or the treating oncologist felt the drug should be prioritized over other therapeutic options.

2) Accepted and deferred - The targeted agent was FDA-approved, had published phase I safety data in children, and/or the treating oncologist felt comfortable prescribing the treatment but chose not to do so in favor of another option for which there was more published evidence of efficacy.

3) Declined - The targeted agent was either not FDA-approved, lacked phase I safety data in children, or existing clinical trial evidence suggested a lack of efficacy.

For assessing the patient-level value of CARE IMPACT findings, the finding deemed most promising for each patient by the treating clinician was used for further categorization.

#### **Determining analysis turnaround time**

Turnaround times were calculated for different steps in the clinical registry study workflow (Appendix Table A9). For each sample different timepoints were collected including date of tumor sample collection, date sample was shipped to Covance for sequencing, date sample was received at Covance, date RNA-Seq files were sent to UCSC Treehouse, date automated Treehouse analysis process completed, date Treehouse hosted an internal mock clinical genomics tumor board meeting, date of final clinical genomics tumor board meeting with UCSC and Stanford. Of note, in some cases a banked sample was used for analysis resulting in a large time from sample collection to being sent to Covance for sequencing.

#### **Data analysis and figures**

Calculations were performed and figures generated with R and RStudio using the following packages: tidyverse, colorspace, cowplot, ggVennDiagram, ggforce, ggrepel, gridExtra, haven, janitor, jsonlite, kableExtra, khroma, knitr, networkD3, RColorBrewer, redcapAPI, UpSetR, webshot.

### **Statement of Data Analysis**

Throughout the duration of this registry study the following individuals from the UC Santa Cruz Treehouse team were involved in data analysis: Yvonne Vasquez, Lauren Sanders, Holly Beale, Ellen Kephart, and Geoffry Lyle.

### **Supplemental Results**

#### **Patient cases where Treehouse findings were prioritized and implemented**

##### **A) Myoepithelial Carcinoma**

A young male (TH34_1352_S01) was diagnosed and treated for non-metastatic myoepithelial carcinoma of the liver. After 26 months of ifosfamide/doxorubicin chemotherapy and complete tumor resection he developed bilateral pulmonary metastases. Molecular testing of the pulmonary metastasis found INI-1 deficiency (Appendix Table A4). RNA-Seq from the same metastatic lung nodule underwent CARE, which found multiple outlier RTKs (FGFR1, FGFR2, PDGFRA), consistent with pan-disease enrichment in FGFR and PDGF pathway signaling, all of which could be targeted with pazopanib (https://www.selleckchem.com/products/pazopanib.html). CARE also identified CCND2 as a pan-disease outlier with pathway support, consistent with the role of SMARCB-1 as a repressor of the cell cycle (Appendix Table A3). This abnormality is targetable by ribociclib.^12^ Therefore, Treehouse analysis nominated both pazopanib and ribociclib for this patient.

After ribociclib treatment, surgical resection and ribociclib maintenance treatment the patient shows no evidence of disease. This case is described in detail in a separate manuscript.

##### **B) Gastrointestinal Stromal Tumor**

A teenage male (TH34_1349_S01/S02) with a germline mutation in *SDHC* (Appendix Table A4) developed recurrent wild-type gastrointestinal stromal tumor (GIST) in the posterior stomach wall with regional nodal and liver metastases nearly 7 years after undergoing wide resection alone. Further tumor progression after 15 months of imatinib therapy prompted complete resection of the gastric mass and enlarged regional lymph nodes and subtotal resection of unresectable liver metastases; pathology confirmed the presence of recurrent GIST with nodal and liver involvement.

RNA-Seq data from the gastric mass and from a liver metastasis were analyzed, and findings in both samples were prioritized as treatment targets, since any treatment based on them could target both sites of the disease. In the gastric sample only, CARE identified *IGF2* as a pan-cancer overexpression outlier with pan-cancer and pan-disease pathway enrichment in the hallmark KRAS signaling pathway. In the liver sample only, *PTCH1* was identified as a pan-cancer and pan-disease overexpression outlier, accompanied by pan-cancer pathway enrichment of the hallmark Hedgehog signaling pathway (Appendix Table A3).

In both samples, CARE found *KIT* and *ETV1* as pan-cancer and pan-disease overexpression outliers. The receptor tyrosine kinase *KIT* activates the RAS/RAF/MEK pathway,^13^ consistent with pan-cancer and pan-disease pathway enrichment in the hallmark KRAS signaling pathway in both samples. This pathway activates transcription factor *ETV1*, leading to cell invasiveness and metastasis as well as transcription of *KIT*.^14^ Most GIST tumors harbor an activating *KIT* mutation which makes them sensitive to kinase inhibitors imatinib and sunitinib.^15^ This tumor did not have a *KIT* mutation but still displayed overexpression at the RNA level, indicating that *KIT* signaling may be a tumor driver. Several studies have shown benefit of sunitinib treatment for GIST patients with wild-type *KIT,*^16–19^ suggesting that *KIT* RNA overexpression may be another predictive biomarker of response to KIT inhibitors, especially in the setting of wild-type *KIT*.

The patient’s therapy was switched to sunitinib. This line of treatment was consistent with the Treehouse analysis, although sunitinib is also standard of care second-line therapy for GIST^20,21^. MRI imaging 37 months after starting sunitinib showed stable disease.

##### **C) Osteosarcoma**

In one case (TH34_1456_S02) a therapy was chosen which targets a Treehouse analysis finding, but this therapy was chosen for a different reason. A teenage male patient had been diagnosed with osteosarcoma metastatic to the lung. CARE analysis of RNA-Seq data from a lung metastasis identified *KDR/VEGFR2* as a pan-cancer and pan-disease up-outlier with pathway support (Appendix Table A3), consistent with *VEGFA* amplification found by STAMP panel (Appendix Table A4). The clinician chose treatment with cabozantinib, an RTK inhibitor, because it has relatively high rates of disease control in recurrent osteosarcoma and is well-tolerated.^22^ The patient had a partial response after 2 months and continued stable disease for an additional 12 months thereafter. Although this treatment was chosen based on prior evidence of efficacy in osteosarcoma, cabozantinib is an RTK inhibitor with high affinity for *KDR/VEGFR2*, and the patient’s response may be related to the outlier expression of *KDR/VEGFR2*.

##### **D) Posterior Fossa Ependymoma**

A young female (TH34_1381_S01) with posterior fossa anaplastic ependymoma with loss of H3K27 trimethylation by immunohistochemistry underwent resection of a frontal lobe metastasis after past treatment with surgery and radiotherapy for the primary tumor, a local recurrence, and a metachronous metastatic recurrence in the same area of the frontal lobe. Loss of H3K27 trimethylation is a hallmark of posterior fossa ependymoma group A (PFA), and is associated with very poor prognosis.^23,24^ Foundation Medicine reported amplification of *IKBKE, MCL1,* and *NTRK1* in the tumor (Appendix Table A4). Additional CyberKnife radiotherapy and 10 cycles of oral etoposide were given. Two months after discontinuing etoposide, disease progression was observed in the posterior fossa.

CARE analysis of the frontal lobe metastasis identified *VEGFA* as a pan-cancer and pan-disease overexpression outlier supported by pan-disease enrichment of the Hallmark Hypoxia pathway (Appendix Table A3). *ERBB2* and *PARP1* were also identified as pan-disease up-outliers. *ERBB2* (*NEU*, *HER2*) encodes a member of the epidermal growth factor (EGF) receptor family of receptor tyrosine kinases. A multicenter Phase II clinical trial of *HER2* inhibitor lapatinib showed modest activity against brain tumor metastases in *HER2*-positive breast cancer patients previously treated with trastuzumab.^25^ In addition, a single group phase II study showed that the combination of lapatinib and capecitabine is active as first-line treatment of brain metastases from *HER2*-positive breast cancer.^26^

The patient was enrolled on a clinical trial of oral neratinib, a HER2 inhibitor. An MRI after 2 cycles showed possible pseudo progression, but an MRI after 3 cycles showed definite tumor progression, so the patient was removed from the study and the family elected no further cancer-directed therapy.

##### **E) Embryonal Rhabdomyosarcoma**

A young female (TH34_2351_S01) with left neck embryonal rhabdomyosarcoma metastatic to multiple bones was treated with multiagent chemotherapy, delayed resection of the primary tumor and radiotherapy. A left neck nodal recurrence 4 years after therapy completion was treated with vinorelbine, cyclophosphamide, and temsirolimus, surgery, and radiotherapy. Three months after therapy completion, a nasopharyngeal recurrence was identified and treated with vincristine, irinotecan, and temozolomide. A subsequent biopsy of the nasopharyngeal recurrence was sent for RNA sequencing and then to Treehouse for analysis.

While awaiting CARE results, the patient was started on pazopanib, because it is FDA-approved for recurrent soft tissue sarcomas in adults and some preclinical data suggests a benefit in pediatric patients.^27,28^ CARE found only one targetable overexpression outlier: *HMOX1* was a pan-disease outlier with pathway support in the PID HIF1 TF Pathway and Hallmark P53 pathway (Appendix Table A3, Figure A4). Additionally, Treehouse RNA variant calling identified an activating *NRAS* G12D mutation (alt/ref 255/103). Because *NRAS* is an upstream activator of *MAP2K1/MEK*, we investigated the expression of *MAP2K1/MEK* and found that it had expression above the 95th percentile in the Treehouse compendium but was not an overexpression outlier.

In the absence of clinical improvement on pazopanib, the patient was switched to trametinib, a *MEK* inhibitor, but 20 days later trametinib was stopped due to symptomatic local progression in the face, for which the patient received palliative RT. The lack of response to trametinib may be attributable to the fact that our assay focuses on targetable overexpression outliers, but *MEK* did not have outlier expression.

#### **Patient cases where Treehouse findings were prioritized but not implemented**

A teenage male (TH34_1240_S01) with pulmonary metastatic Ewing sarcoma developed progressive pulmonary metastases along with nodal and bony metastases while receiving up-front standard chemotherapy and after undergoing delayed wide resection of the primary left scapular tumor. Further disease progression in the lungs and lymph nodes, along with soft tissue and liver metastases were observed despite treatment with several salvage chemotherapy regimens. Following Treehouse findings that MYC was a pan-disease overexpression outlier with pathway support (Appendix Table A3), the patient would have been placed on a phase I clinical trial of a MYC-targeted drug, but the patient was too ill to qualify for the study and he died without receiving any further systemic therapy.

In a second case, a young male patient (TH34_1379_S01) was diagnosed with hepatoblastoma/transitional cell carcinoma of the liver, with later recurrence of disease. The Treehouse analysis identified *FGFR1* as a pan-disease overexpression outlier, supported by pathway enrichment in the Hallmark Angiogenesis Pathway (Appendix Table A3). FGFR proteins can be targeted by pazopanib, an RTK inhibitor. The patient received a cycle of docetaxel and the clinician planned to give a second cycle of docetaxel along with pazopanib, but the patient died before receiving further therapy.

A teenage male (TH34_1380_S01) was diagnosed with embryonal rhabdomyosarcoma with multiple metastases. Several genomic alterations were noted by Foundation Medicine and STAMP (Appendix Table A4). CARE identified *NOTCH3* (therapy: tarextumab) as a pan-cancer and pan-disease up-outlier with pathway support, and *IGF2* as a pan-cancer up-outlier (therapy: ganitumab)(Appendix Table A3). The clinician elected to give pazopanib instead because it is FDA-approved for recurrent sarcomas in adults and there is some evidence of clinical utility in rhabdomyosarcoma. However, disease progression was observed after 2 cycles and palliative RT was given due to pain and spinal cord compression. During RT, the clinician initiated a compassionate use IND for the NOTCH inhibitor nirogacestat. However, oral etoposide was subsequently started since the patient’s mother did not want to wait any longer for systemic therapy, and etoposide is known to be an active agent in rhabdomyosarcoma. He was subsequently transitioned to hospice care.

A male patient (TH34_1455_S01) with relapsed ependymoma would have been placed on a neratinib phase II study following Treehouse findings that *ERBB2* was a pan-disease overexpression outlier (Appendix Table A3), but he was ineligible for the trial due to elevated liver function tests.

A young adult male patient (TH34_2292_S01) was diagnosed with undifferentiated round cell sarcoma with brain metastases. Foundation Medicine reported CIC rearrangement, exon 20 (Appendix Table A4). Prior to CARE, the patient was started on trabectedin because there is some evidence that it may be useful in CIC-rearranged soft tissue sarcomas. CARE identified *FLT4* (therapy: pazopanib) as a pan-cancer up-outlier with pathway support, and *VEGFA* (therapy: bevacizumab) as a pan-cancer up-outlier with pathway support (Appendix Table A3). The clinician was interested in the Treehouse findings, but the patient was not able to get either therapy because he was unable to tolerate any oral medications (pazopanib) and he did not get bevacizumab because he was already on trabectedin by the time the Treehouse results were available. The clinician would have considered bevacizumab therapy after trabectedin, but the patient was never well enough to receive further therapy and died after resection of the brain metastases.

#### **Patient with a rare tumor with no standard therapies**

A teenage female who presented with severe anemia (hemoglobin 4.1 g/dL) underwent partial excision of a necrotic, pedunculated mass extending from the uterus through the cervical os; pathology showed high-grade endometrial stromal sarcoma with YWHAE-NUTM2 gene fusion. Laparoscopic total hysterectomy and bilateral pelvic lymph node dissection confirmed tumor extension to the parametrium and lymph node metastases. Computed tomography imaging of the chest showed 3 small, solid bilateral pulmonary nodules suggestive of metastases. In the absence of a clear standard of care, particularly in a pediatric patient, dose-intensive ifosfamide/doxorubicin chemotherapy, and external beam radiotherapy 48.6 Gy to the whole pelvis were given. Due to the anticipated very poor prognosis, a portion of the primary tumor resection specimen was sent to Treehouse for CARE analysis.

CARE analysis of RNA-Seq data from a primary tumor resection identified the estrogen receptor *ESR1* as a pan-cancer and pan-disease overexpression outlier (Appendix Table A3). Downstream targets of ESR1, CCND1 and CDK4,^29^ were found to be highly expressed in the sample and CDK4/6 inhibitor palbociclib was recommended as a potential therapy. CARE also identified elevated expression of *MAPK11*, which is known to lead to increases in cell survival,^30^ and *ABCB1*, which was identified as a pan-disease overexpression outlier. Based on *ABCB1* being known to increase resistance to chemotherapy,^31,32^ regorafenib and ulixertinib were recommended for their known ability to overcome *ABCB1*-mediated chemotherapeutic drug resistance.^33,34^ In this case, the clinician chose to defer all Treehouse findings while the patient received standard chemotherapy and radiotherapy. The patient currently remains free of disease recurrence 22 months from therapy completion.

#### **Turnaround time**

The median time from collection of a patient tissue sample to submission of the CARE IMPACT report to the treating oncologist was 20 days (range 8-38 days)(Appendix Table A9). The turnaround time remained consistent throughout the study. In general, overall turnaround times remained consistent throughout the study regardless of disease type.

**Adding more datasets to comparator cohorts improves the specificity**

The size and the quality of RNA sequencing datasets in comparator cohorts affects outlier results. As part of our standard process, our data compendia are updated regularly with new datasets. We also routinely perform quality control analysis, including review of the annotations that accompany datasets, and investigations for possible batch effects. We discovered annotation errors affecting 20 of the 27 ependymomas in version 8; these datasets were removed in the subsequent versions of the compendium. By compendium version v11, we had also added 95 high quality ependymoma datasets from 5 studies. Consequently, while ERBB2 was detected as a pan-disease outlier by CARE when the TH34_1381_S01 dataset was analyzed using the Treehouse compendium version 8 (v8), this outlier was not detected against the updated compendium version 11 (v11).

The 6 datasets most highly correlated to TH34_1381_S01 in the v8 compendium included ependymoma, glioblastoma multiforme and glioma. All correlations were above our required threshold^21^ of 0.875 for that compendium, with the highest correlation being 0.90. The "diseases of the top 6 most correlated datasets" comparator cohort consisted of datasets originating from the three diseases, and TH34_1381_S01's ERBB2 expression exceeded the outlier threshold. It also exceeded the threshold for the "1st and 2nd degree correlated samples" cohort, making it a consensus pan-disease outlier. The patient was treated with an ERBB2 inhibitor on a clinical trial but experienced disease progression.

In contrast, all 6 of the most correlated datasets identified in the v11 compendium were ependymoma, and the lowest correlated was 0.92. Consequently, the "diseases of the top 6 most correlated datasets" comparator cohort in v11 included only ependymomas. TH34_1381_S01's ERBB2 expression did not exceed the outlier threshold of the cohort. The new ependymomas indicated that TH34_1381_S01's ERBB2 expression was not exceptional for ependymomas. Although the expression also exceeded the threshold for the "1st and 2nd-degree correlated samples" cohort, it was not a consensus pan-disease outlier and was not reported in Table 1 or Figure 3. This case illustrates the importance of increasing the size of the comparator cohort with high-quality datasets.

### **Data Acknowledgment**

Use of data available in public repositories allowed us to increase the number of pediatric cancer patients and pediatric cancer types included in our reference compendium; we gratefully acknowledge the data providers listed in Appendix Table A5 in the manner they have specified.

Use of data from partners and available in public repositories allowed us to increase the number of pediatric cancer patients and pediatric cancer types included in our reference compendium, and we gratefully acknowledge the following data providers in the manner they have specified:

**Tumor Compendium v11 Public PolyA (April 2020)**

This research was conducted using data made available by the following institutions:

British Columbia Cancer Agency

The Children’s Brain Tumor Tissue Consortium

The Hospital for Sick Children

Stanford University

University of Calgary

University of California, San Francisco

International Cancer Genome Consortium. The following ICGC datasets were used in this work: EGAD00001000158, EGAD00001001620, EGAD00001000328, EGAD00001000648, EGAD00001000617, EGAD00001000826 and EGAD00001000356.

St. Jude Children’s Research Hospital – Washington University Pediatric Cancer Genome Project; the following datasets were obtained via EGA ( https://www.ebi.ac.uk/ega/ ) and were used by permission: EGAD00001001098: Andersson AK, Ma J, Wang J, et al. The landscape of somatic mutations in infant MLL-rearranged acute lymphoblastic leukemias. Nat Genet. 2015;47(4):330-7. (PMC4553269). EGAD00001001666: Qaddoumi I, Orisme W, Wen J, et al. Genetic alterations in uncommon low-grade neuroepithelial tumors: BRAF, FGFR1, and MYB mutations occur at high frequency and align with morphology. Acta Neuropathol. 2016;131(6):833-45. (PMID 26810070). EGAD00001002680: Pinto EM, Chen X, Easton J, et al. Genomic landscape of paediatric adrenocortical tumours. Nat Commun. 2015;6:6302. (PMC4352712). Dataset SJC-DS-1001 was accessed and processed with permission from St. Jude Cloud (https://www.stjude.cloud) – a publicly accessible pediatric genomic data resource requiring approval for controlled data access:

Chen, X, et al. Targeting oxidative stress in embryonal rhabdomyosarcoma. Cancer Cell. 2013 Dec 9;24(6):710-24.

Gruber TA, et al. An Inv(16)(p13.3q24.3)-encoded CBFA2T3-GLIS2 fusion protein defines an aggressive subtype of pediatric acute megakaryoblastic leukemia. Cancer Cell. 2012; 22(5):683-697.

Holmfeldt L, et al. The genetic landscape of hypodiploid acute lymphoblastic leukemia. Nat. Genet. 2013.

Robinson G, et al. Novel mutations target distinct subgroups of medulloblastoma. Nature. 2012; 488(7409):43-48.

Zhang J, et al. Whole-genome sequencing identifies genetic alterations in pediatric low-grade gliomas. Nat. Genet. 2013; 45(6):602-12.

Zhang J, et al. A novel retinoblastoma therapy from genomic and epigenetic analyses. Nature. 2012; 481(7381):329-334.

Zhang J, et al. The genetic basis of early T-cell precursor acute lymphoblastic leukaemia. Nature. 2012; 481(7380):157-163.

Provider(s) of the data under accession phs000178.v10.p8 at dbGap. The Cancer Genome Atlas Research Network, National Cancer Institute and National Human Genome Research Institute, Bethesda, MD, USA. The results published here are in part based upon data generated by The Cancer Genome Atlas managed by the NCI and NHGRI. Information about TCGA can be found at http://cancergenome.nih.gov/.

Provider(s) of the data under accession phs000178.v10.p8 at dbGap: The Therapeutically Applicable Research to Generate Effective Treatments (TARGET) initiative managed by the NCI. The data used for this analysis are available at dbGap under accession phs000218. Information about TARGET can be found at http://ocg.cancer.gov/programs/target.

Children's Oncology Group (COG; Tissues for TARGET are collected as part of COG clinical and biological protocols)

Peter C. Adamson, MD. Children's Hospital of Philadelphia, Philadelphia, PA, USA

Principal Investigator (ALL Project Team)

Stephen P. Hunger, MD. University of Colorado Cancer Center, Denver, CO, USA

Principal Investigators (AML Project Team)

Soheil Meshinchi, MD, PhD. Fred Hutchinson Cancer Research Center, Seattle, WA, USA

Robert Arceci, MD, PhD. Children's Hospital, Phoenix, AZ, USA

Principal Investigator (NBL Project Team)

John M. Maris, MD. Children's Hospital of Philadelphia, Philadelphia, PA, USA

Robert Seeger, MD. Children's Hospital of Los Angeles, Los Angeles, CA, USA

Javed Khan, MD. National Cancer Institute, National Institutes of Health, Bethesda, MD, USA

Principal Investigator (OS Project Team)

Ching Lau, MD, PhD. Texas Children's Hospital, Houston, TX, USA

Paul Meltzer, MD, PhD. National Cancer Institute, National Institutes of Health, Bethesda, MD, USA

Principal Investigator (Kidney Project Teams - WT, CCSK, RT)

Elizabeth J. Perlman, MD. Ann and Robert H. Lurie Children's Hospital of Chicago, Chicago, IL, USA

Principal Investigator (Cell Lines and Xenografts - PPTP)

Peter Houghton, PhD. The Research Institute at Nationwide Children's Hospital, Columbus, OH, USA

Provider(s) of the data under accession phs000699.v1.p1 at dbGap: Todd Golub. Dana Farber Cancer Institute, Boston, MA, USA. This work was conducted as part of the Slim Initiative for Genomic Medicine in the Americas (SIGMA), a project funded by the Carlos Slim Health Institute in Mexico.

Provider(s) of the data under accession phs000900.v1.p1 at dbGap: Michelle Monje, Stanford University, Stanford, CA, USA. We thank the many patients and families who selflessly contributed to this study through tissue donations from surgery or autopsy and Amar Gajjar, for his guidance and vision throughout this study. We also thank Darren Hargrave, James Olson and Sarah Leary for selection of V.2 chemical screen agents. We are grateful for the critical questions and comments by Simone Cheetham and Nadim Nsouli. We also acknowledge important comments by other DIPG Preclinical Consortium member Oren Becher. We thank Gerald Grant for assistance in developing rodent CED techniques. Short read sequencing was performed by the OHSU Massively Parallel Sequencing Shared Resource. The first paper using this data was: Grasso CS, Tang Y, Truffaux N, et al. Functionally defined therapeutic targets in diffuse intrinsic pontine glioma. Nat Med. 2015;21(6):555-9. (PMID 25939062).

Provider(s) of the data under accession phs000720.v2.p1 at dbGap: Javed Khan, MD. National Institutes of Health, Bethesda, MD, USA. The authors thank the Children's Oncology Group Soft Tissue Sarcoma Committee and the BioPathology Center, for their careful collection of clinical samples. This research was supported by the Intramural Research Program of the National Institute of Health and National Cancer Institute.

Provider(s) of the data under accession phs000768.v2.p1 at dbGap. Javed Khan, MD. National Institutes of Health, Bethesda, MD, USA. This research was supported by the Intramural Research Program of the National Institute of Health and National Cancer Institute. The datasets have been accessed through the NIH database for Genotypes and Phenotypes (dbGaP) under accession # phs000768.v1.p1.

Provider(s) of the data under accession phs000673.v2.p1 at dbGap. Arul Chinnaiyan, MD PhD. Michigan Center for Translational Pathology, University of Michigan, MI, USA. The results published here are in whole or part based upon data generated by the Clinical Sequencing Exploratory Research (CSER) consortium established by the NHGRI. Funding support was provided through cooperative agreements with the NHGRI and NCI through grant numbers U01 HG006508 (Exploring Cancer Medicine for Sarcoma and Rare Cancers). Information about CSER and the investigators and institutions who comprise the CSER consortium can be found at http://www.genome.gov/27546194

Provider(s) of the data under accession SRP040454 at SRA. Dataset was made available by Memorial Sloan Kettering Cancer Center.

### **Supplemental Figures**

#### **Figure A1. Clinical utility of CARE IMPACT findings in 33 patients**

The findings are arranged on the y-axis and grouped into “Accepted” and “Declined” categories as described in Supplemental Methods. The details of the findings are described in Supplemental Results.

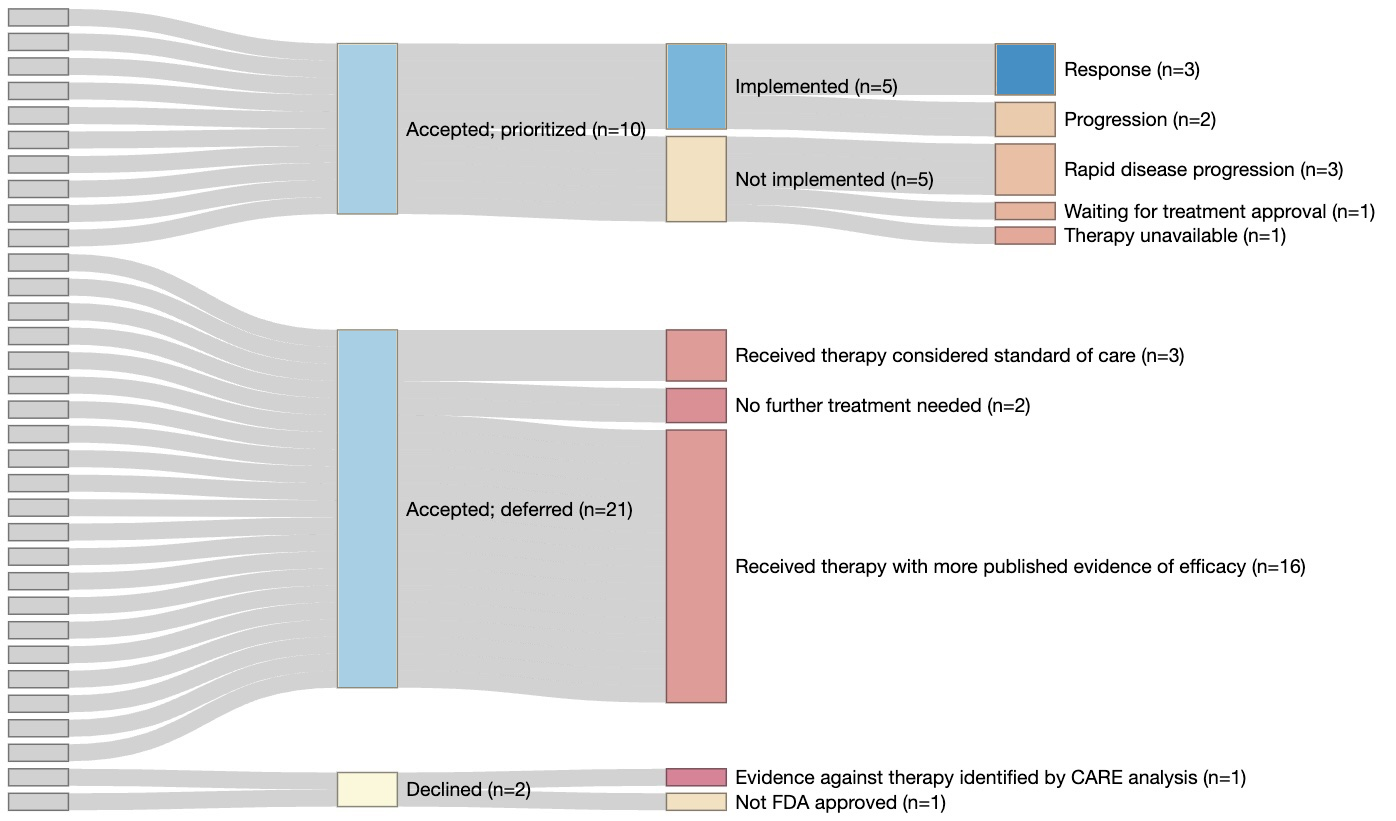

#### **Figure A2. Outliers detected by different comparative cohorts**

The outlier gene is shown on the y-axis, while patient RNA-Seq sample ID’s are shown on the x-axis. The colors indicate the comparator cohort used to identify the outlier gene.

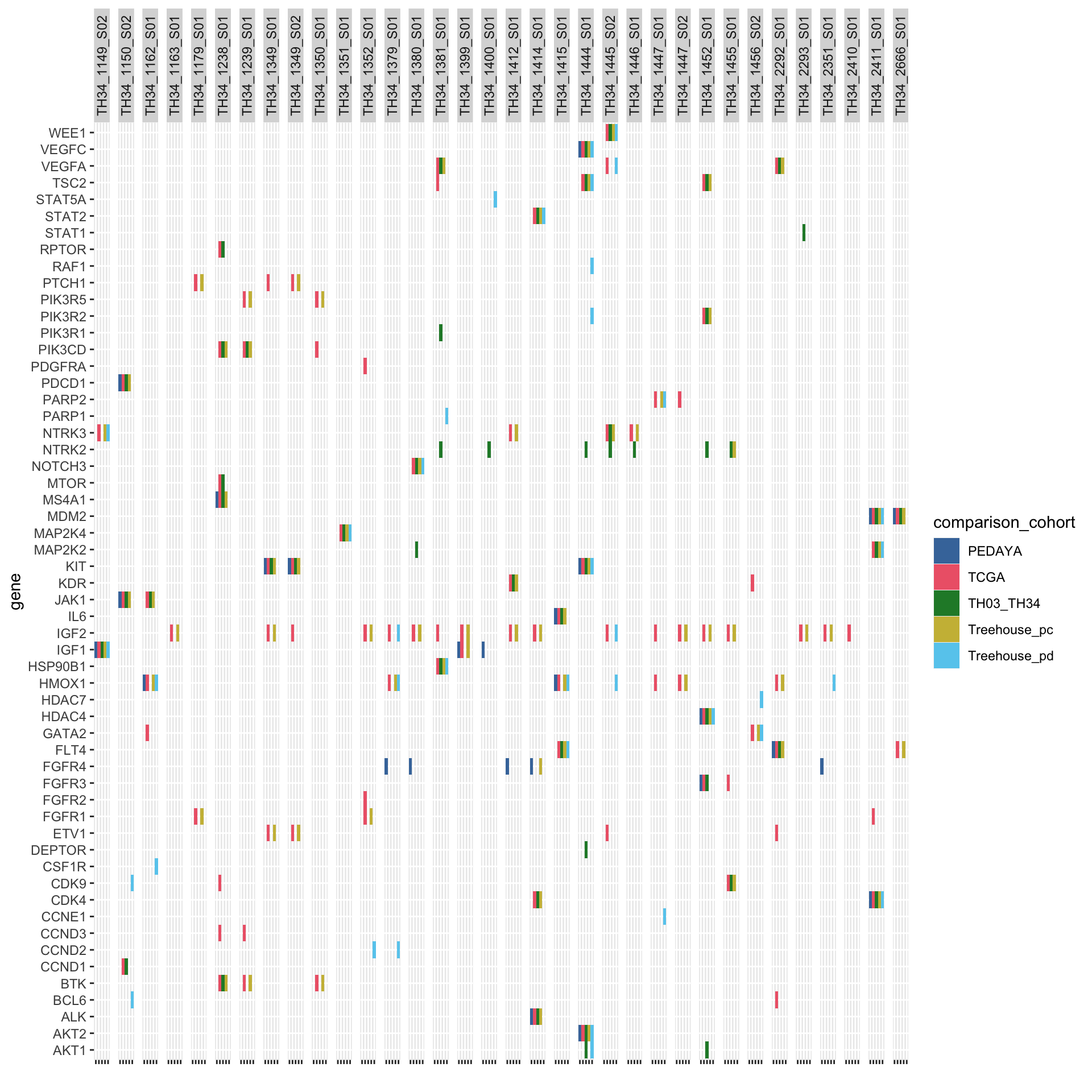

#### **Figure A3. Outliers detected relative to cohorts**

The total number of outliers detected relative to each cohort is displayed on the horizontal bars on the left of the cohort name. Vertical bars depict the number of outliers detected relative to each combination of cohorts indicated by black points. The largest set consists of 27 outliers detected relative to both the Treehouse pan-cancer cohort and TCGA. Combinations yielding no outliers are omitted.

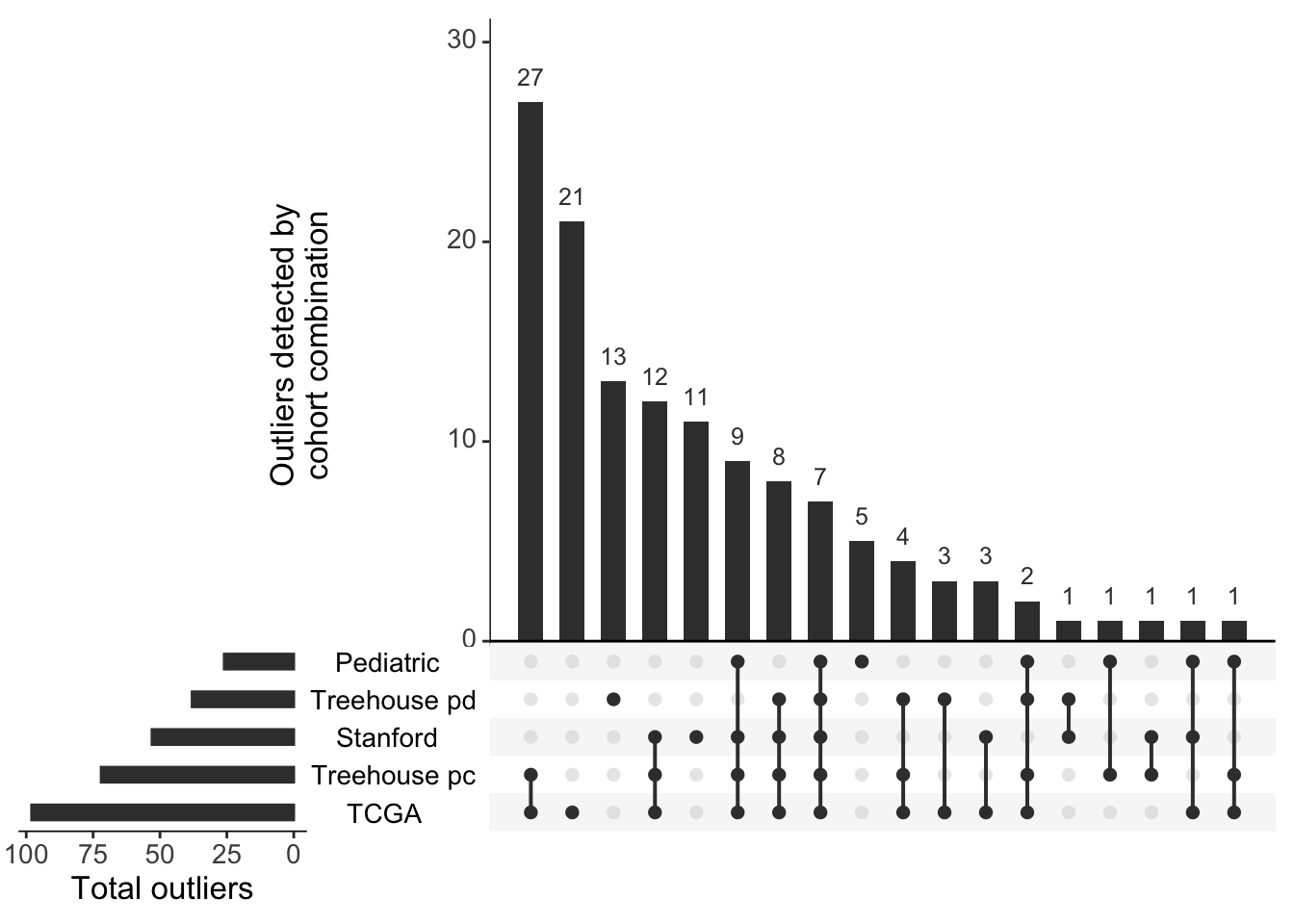

### **Supplemental Tables**

#### **Table A1. Patient demographics and clinical information**

| **Patient ID** | **Age at Diagnosis Range (years)** | **Sex** | **Race** | **Ethnicity** | **Diagnosis** | **Disease Group** | **Clinical_Status** |
| --- | --- | --- | --- | --- | --- | --- | --- |
| TH34_1149 | 0-5 | male | White | Not Hispanic or Latino | INI-deficient soft tissue sarcoma NOS | Sarcoma other (all other types) | Recurrence or relapse of disease |
| TH34_1150 | 16-20 | male | Asian | Not Hispanic or Latino | ewing sarcoma | Sarcoma bone (osteo and Ewing) | Recurrence or relapse of disease |
| TH34_1162 | 16-20 | female | White | Not Hispanic or Latino | ewing sarcoma | Sarcoma bone (osteo and Ewing) | Recurrence or relapse of disease |
| TH34_1163 | 16-20 | male | Asian | Not Hispanic or Latino | embryonal rhabdomyosarcoma | Sarcoma other (all other types) | Recurrence or relapse of disease |
| TH34_1179 | 21-25 | female | White | Not Hispanic or Latino | colon adenocarcinoma | Other | Refractory disease |
| TH34_1238 | 0-5 | female | Other | Hispanic or Latino | acute lymphoblastic leukemia | Hematopoietic | Refractory disease |
| TH34_1239 | 0-5 | female | White | Not Hispanic or Latino | acute myeloid leukemia | Hematopoietic | Refractory disease |
| TH34_1240 | 16-20 | male | White | Not Hispanic or Latino | ewing sarcoma | Sarcoma bone (osteo and Ewing) | Recurrence or relapse of disease |
| TH34_1349 | 11-15 | male | Other | Hispanic or Latino | gastrointestinal stromal tumor | Sarcoma other (all other types) | Recurrence or relapse of disease |
| TH34_1350 | 0-5 | female | Other | Hispanic or Latino | juvenile myelomonocytic leukemia | Hematopoietic | Recurrence or relapse of disease |
| TH34_1351 | 6-10 | female | Other | Hispanic or Latino | osteosarcoma | Sarcoma bone (osteo and Ewing) | Recurrence or relapse of disease |
| TH34_1352 | 0-5 | male | Other | Hispanic or Latino | myoepithelial carcinoma | Sarcoma other (all other types) | Recurrence or relapse of disease |
| TH34_1379 | 0-5 | male | Other | Hispanic or Latino | hepatoblastoma | Liver | Recurrence or relapse of disease |
| TH34_1380 | 11-15 | male | Other | Hispanic or Latino | embryonal rhabdomyosarcoma | Sarcoma other (all other types) | Recurrence or relapse of disease |
| TH34_1381 | 0-5 | female | White | Not Hispanic or Latino | ependymoma | CNS | Recurrence or relapse of disease |
| TH34_1399 | 11-15 | male | White | Not Hispanic or Latino | desmoplastic small round cell tumor | Sarcoma other (all other types) | Recurrence or relapse of disease |
| TH34_1400 | 16-20 | female | White | Not Hispanic or Latino | pleomorphic myxoid liposarcoma | Sarcoma other (all other types) | Recurrence or relapse of disease |
| TH34_1412 | 21-25 | male | Asian | Not Hispanic or Latino | desmoplastic small round cell tumor | Sarcoma other (all other types) | Recurrence or relapse of disease |
| TH34_1414 | 16-20 | female | Asian | Not Hispanic or Latino | alveolar rhabdomyosarcoma | Sarcoma other (all other types) | Recurrence or relapse of disease |
| TH34_1415 | 16-20 | female | Asian | Not Hispanic or Latino | undifferentiated sarcoma nos | Sarcoma other (all other types) | Recurrence or relapse of disease |
| TH34_1444 | 0-5 | male | Other | Hispanic or Latino | follicular neoplasm | Endocrine tumor (adrenal, thyroid) | Recurrence or relapse of disease |
| TH34_1445 | 6-10 | female | Other | Hispanic or Latino | glioma | CNS | Recurrence or relapse of disease |
| TH34_1446 | 0-5 | female | White | Not Hispanic or Latino | ganglioglioma | CNS | Recurrence or relapse of disease |
| TH34_1447 | 0-5 | female | Asian | Not Hispanic or Latino | undifferentiated hepatic sarcoma | Sarcoma other (all other types) | Recurrence or relapse of disease |
| TH34_1452 | 0-5 | male | Other | Hispanic or Latino | ependymoma | CNS | Recurrence or relapse of disease |
| TH34_1455 | 0-5 | male | Other | Hispanic or Latino | ependymoma | CNS | Recurrence or relapse of disease |
| TH34_1456 | 11-15 | male | Other | Hispanic or Latino | osteosarcoma | Sarcoma bone (osteo and Ewing) | Recurrence or relapse of disease |
| TH34_2292 | 21-25 | male | Other | Hispanic or Latino | undifferentiated sarcoma nos | Sarcoma other (all other types) | Recurrence or relapse of disease |
| TH34_2293 | 0-5 | male | White | Not Hispanic or Latino | sclerosing epithelioid fibrosarcoma | Sarcoma other (all other types) | Recurrence or relapse of disease |
| TH34_2351 | 0-5 | female | Asian | Not Hispanic or Latino | embryonal rhabdomyosarcoma | Sarcoma other (all other types) | Recurrence or relapse of disease |
| TH34_2410 | 16-20 | female | Other | Hispanic or Latino | endometrial stromal sarcoma | Sarcoma other (all other types) | Initial diagnosis of disease |
| TH34_2411 | 16-20 | male | Other | Not Hispanic or Latino | osteosarcoma | Sarcoma bone (osteo and Ewing) | Recurrence or relapse of disease |
| TH34_2666 | 21-25 | male | White | Not Hispanic or Latino | soft tissue sarcoma | Sarcoma other (all other types) | Recurrence or relapse of disease |

#### **Table A2. Directly and indirectly actionable genes used to prioritize gene expression outlier findings**

| **Gene** | **Category** | **Druggability** | **FDA approved drug** | **Non-FDA approved drugs** |
| --- | --- | --- | --- | --- |
| AURKA | Aurora Kinases | directly druggable gene/gene family |  | alisertib |
| AURKB | Aurora Kinases | directly druggable gene/gene family |  | alisertib |
| AURKC | Aurora Kinases | directly druggable gene/gene family |  | alisertib |
| BTK | B-cell Receptor Signaling | directly druggable gene/gene family | ibrutinib |  |
| MS4A1 | B-cell Receptor Signaling | directly druggable gene/gene family | ibritumomab tiuxetan, obinutuzumab, ofatumumab, rituximab |  |
| BCL2 | BCL2/MDM2 | directly druggable gene/pathway | venetoclax |  |
| BCL6 | BCL2/MDM2 | indirectly actionable gene (BCL2) |  |  |
| MDM2 | BCL2/MDM2 | indirectly actionable gene (BCL2) |  | APG-115, ALRN-6924 |
| CDK4 | Cell Cycle | directly druggable gene/pathway | palbociclib, ribociclib |  |
| CDK6 | Cell Cycle | directly druggable gene/pathway | palbociclib, ribociclib |  |
| CCND1 | Cell Cycle | indirectly actionable gene (CDK) |  |  |
| CCND2 | Cell Cycle | indirectly actionable gene (CDK) |  | ribociclib |
| CCND3 | Cell Cycle | indirectly actionable gene (CDK) |  |  |
| CCNE1 | Cell Cycle | indirectly actionable gene (CDK) |  | dinaciclib |
| CDK2 | Cell Cycle | directly druggable gene/pathway |  | dinaciclib |
| PARP1 | DNA Repair | directly druggable gene/gene family | olaparib, niraparib |  |
| PARP2 | DNA Repair | directly druggable gene/gene family | olaparib, niraparib |  |
| HDAC4 | HDAC | directly druggable gene/gene family | belinostat, panobinostat, valproic acid, vorinostat |  |
| HDAC7 | HDAC | directly druggable gene/gene family | belinostat, panobinostat, valproic acid, vorinostat |  |
| HSP90AA1 | Heat Shock Proteins | directly druggable gene/pathway |  |  |
| HSP90AA2P | Heat Shock Proteins | directly druggable gene/gene family |  |  |
| HSP90N | Heat Shock Proteins | directly druggable gene/gene family |  |  |
| HSP90AA2 | Heat Shock Proteins | directly druggable gene/gene family |  |  |
| HSP90AA6P | Heat Shock Proteins | directly druggable gene/gene family |  |  |
| HSP90AA3P | Heat Shock Proteins | directly druggable gene/gene family |  |  |
| HSP90AA4P | Heat Shock Proteins | directly druggable gene/gene family |  |  |
| HSP90AA5P | Heat Shock Proteins | directly druggable gene/gene family |  |  |
| HSP90AB1 | Heat Shock Proteins | directly druggable gene/gene family |  |  |
| HSP90AB3P | Heat Shock Proteins | directly druggable gene/gene family |  |  |
| HSP90AB2P | Heat Shock Proteins | directly druggable gene/gene family |  |  |
| HSP90AB6P | Heat Shock Proteins | directly druggable gene/gene family |  |  |
| HSP90AB4P | Heat Shock Proteins | directly druggable gene/gene family |  |  |
| HSP90AB5P | Heat Shock Proteins | directly druggable gene/gene family |  |  |
| HSP90B1 | Heat Shock Proteins | directly druggable gene/gene family |  |  |
| HSP90B2P | Heat Shock Proteins | directly druggable gene/gene family |  |  |
| HSP90B3P | Heat Shock Proteins | directly druggable gene/gene family |  |  |
| HMOX1 | HIF1A Pathway | directly druggable gene/pathway |  |  |
| PDCD1 | Immune Checkpoint | directly druggable gene/gene family | nivolumab, pembrolizumab, atezolizumab |  |
| JAK1 | JAK/STAT | directly druggable gene/pathway | ruxolitinib, tofacitinib |  |
| JAK2 | JAK/STAT | directly druggable gene/pathway | ruxolitinib, tofacitinib |  |
| JAK3 | JAK/STAT | directly druggable gene/pathway | tofacitinib |  |
| STAT1 | JAK/STAT | indirectly actionable gene (JAK) |  |  |
| STAT2 | JAK/STAT | indirectly actionable gene (JAK) |  |  |
| STAT3 | JAK/STAT | directly druggable gene/pathway |  |  |
| STAT5A | JAK/STAT | indirectly actionable gene (JAK) |  | ruxolitinib |
| STAT5B | JAK/STAT | indirectly actionable gene (JAK) |  |  |
| IL6 | JAK/STAT | directly druggable gene/pathway | siltuximab |  |
| IL6R | JAK/STAT | directly druggable gene/pathway |  | tocilizumab |
| MAP2K4 | JNK Pathway | directly druggable gene/pathway |  | vemurafenib |
| NOTCH3 | NOTCH | directly druggable gene/gene family |  | tarextumab, nirogacestat |
| PIK3CA | PI3K/AKT/mTOR | directly druggable gene/pathway | idelalisb, everolimus, sirolimus |  |
| PIK3CB | PI3K/AKT/mTOR | directly druggable gene/pathway | idelalisb, sirolimus |  |
| PIK3CG | PI3K/AKT/mTOR | directly druggable gene/pathway | idelalisib |  |
| PIK3CD | PI3K/AKT/mTOR | directly druggable gene/pathway | idelalisb, everolimus |  |
| PIK3R1 | PI3K/AKT/mTOR | indirectly actionable gene (mTOR) | everolimus |  |
| PIK3R2 | PI3K/AKT/mTOR | indirectly actionable gene (mTOR) | everolimus, sirolimus, temsirolimus |  |
| PIK3R3 | PI3K/AKT/mTOR | indirectly actionable gene (mTOR) | everolimus, sirolimus, temsirolimus |  |
| PIK3R4 | PI3K/AKT/mTOR | indirectly actionable gene (mTOR) | everolimus, sirolimus, temsirolimus |  |
| PIK3R5 | PI3K/AKT/mTOR | indirectly actionable gene (mTOR) | everolimus, sirolimus, temsirolimus |  |
| PIK3R6 | PI3K/AKT/mTOR | indirectly actionable gene (mTOR) | everolimus, sirolimus, temsirolimus |  |
| PIK3C2A | PI3K/AKT/mTOR | indirectly actionable gene (mTOR) | everolimus, sirolimus, temsirolimus |  |
| PIK3C2B | PI3K/AKT/mTOR | indirectly actionable gene (mTOR) | everolimus, sirolimus, temsirolimus |  |
| PIK3C2G | PI3K/AKT/mTOR | indirectly actionable gene (mTOR) | everolimus, sirolimus, temsirolimus |  |
| PIK3C3 | PI3K/AKT/mTOR | indirectly actionable gene (mTOR) | everolimus, sirolimus, temsirolimus |  |
| AKT1 | PI3K/AKT/mTOR | indirectly actionable gene (mTOR) | sirolimus | everolimus, AZD5363 |
| AKT3 | PI3K/AKT/mTOR | indirectly actionable gene (mTOR) | everolimus, sirolimus, temsirolimus |  |
| TSC1 | PI3K/AKT/mTOR | indirectly actionable gene (mTOR) | everolimus, sirolimus, temsirolimus |  |
| TSC2 | PI3K/AKT/mTOR | indirectly actionable gene (mTOR) | everolimus, sirolimus, temsirolimus |  |
| MTOR | PI3K/AKT/mTOR | directly druggable gene/pathway | everolimus, sirolimus, temsirolimus |  |
| RPTOR | PI3K/AKT/mTOR | indirectly actionable gene (mTOR) |  |  |
| MLST8 | PI3K/AKT/mTOR | indirectly actionable gene (mTOR) |  |  |
| DEPTOR | PI3K/AKT/mTOR | indirectly actionable gene (mTOR) |  |  |
| AKT2 | PI3K/AKT/mTOR | directly druggable gene/gene family | everolimus, sirolimus, temsirolimus |  |
| GATA2 | Proteasome | directly druggable gene/gene family | bortezomib |  |
| NRAS | RAS/RAF/MEK | indirectly actionable gene (MEK) | trametinib, selumetinib |  |
| KRAS | RAS/RAF/MEK | indirectly actionable gene (MEK) | trametinib, selumetinib |  |
| HRAS | RAS/RAF/MEK | indirectly actionable gene (MEK) | trametinib, selumetinib |  |
| BRAF | RAS/RAF/MEK | directly druggable gene/pathway | dabrafenib, regorafenib, sorafenib, vemurafenib |  |
| RAF1 | RAS/RAF/MEK | directly druggable gene/pathway | dabrafenib, regorafenib, sorafenib, vemurafenib |  |
| ARAF | RAS/RAF/MEK | indirectly actionable gene (MEK) |  |  |
| MAP2K1 | RAS/RAF/MEK | directly druggable gene/pathway | trametinib, selumetinib |  |
| MAP2K2 | RAS/RAF/MEK | directly druggable gene/pathway | trametinib, selumetinib |  |
| ETV1 | RAS/RAF/MEK | indirectly actionable gene (MEK) |  | trametinib, selumetinib |
| EGFR | Receptor Tyrosine Kinase (RTK) | directly druggable gene/pathway | afatinib, cetuximab, erlotinib, gefitinib, lapatinib, panitumumab, vandetanib |  |
| ERBB2 | Receptor Tyrosine Kinase (RTK) | directly druggable gene/pathway | afatinib, cetuximab, erlotinib, gefitinib, lapatinib, panitumumab, vandetanib |  |
| ERBB3 | Receptor Tyrosine Kinase (RTK) | directly druggable gene/pathway | lapatinib |  |
| ALK | Receptor Tyrosine Kinase (RTK) | directly druggable gene/pathway | ceritinib, crizotinib, lorlatinib |  |
| MET | Receptor Tyrosine Kinase (RTK) | directly druggable gene/pathway | cabozantinib, crizotinib |  |
| ROS1 | Receptor Tyrosine Kinase (RTK) | directly druggable gene/pathway | cabozantinib, crizotinib, ceritinib |  |
| FLT1 | Receptor Tyrosine Kinase (RTK) | directly druggable gene/pathway | axitinib, cabozantinib, nintedanib, pazopanib, regorafenib |  |
| FLT4 | Receptor Tyrosine Kinase (RTK) | directly druggable gene/pathway | axitinib, cabozantinib, nintedanib | pazopanib |
| PDGFRA | Receptor Tyrosine Kinase (RTK) | directly druggable gene/pathway | axitinib, cabozantinib, nintedanib, dasatinib, imatinib | pazopanib |
| PDGFRB | Receptor Tyrosine Kinase (RTK) | directly druggable gene/pathway | axitinib, cabozantinib, nintedanib, pazopanib, regorafenib, sorafenib, sunitinib |  |
| FGFR1 | Receptor Tyrosine Kinase (RTK) | directly druggable gene/pathway | nintedanib, pazopanib, ponatinib, regorafenib, erdafitinib |  |
| FGFR2 | Receptor Tyrosine Kinase (RTK) | directly druggable gene/gene family | nintedanib, ponatinib, regorafenib |  |
| FGFR3 | Receptor Tyrosine Kinase (RTK) | directly druggable gene/gene family | nintedanib, pazopanib, ponatinib |  |
| FGFR4 | Receptor Tyrosine Kinase (RTK) | directly druggable gene/gene family | nintedanib, ponatinib |  |
| NTRK1 | Receptor Tyrosine Kinase (RTK) | directly druggable gene/gene family | cabozantinib, ponatinib |  |
| NTRK2 | Receptor Tyrosine Kinase (RTK) | directly druggable gene/gene family | cabozantinib, ponatinib |  |
| NTRK3 | Receptor Tyrosine Kinase (RTK) | directly druggable gene/gene family | cabozantinib, ponatinib | entrectinib |
| FLT3 | Receptor Tyrosine Kinase (RTK) | directly druggable gene/pathway | cabozantinib, ponatinib, sorafenib, sunitinib |  |
| KDR | Receptor Tyrosine Kinase (RTK) | directly druggable gene/pathway | axitinib, cabozantinib, nintedanib, pazopanib, ponatinib, ramucirumab, regorafenib, sorafenib, sunitinib, vandetanib |  |
| KIT | Receptor Tyrosine Kinase (RTK) | directly druggable gene/pathway | axitinib, cabozantinib, dasatinib, imatinib, nilotinib, pazopanib, ponatinib, regorafenib, sorafenib, sunitinib, midostaurin |  |
| CSF1R | Receptor Tyrosine Kinase (RTK) | directly druggable gene/pathway | pazopanib |  |
| TEK | Receptor Tyrosine Kinase (RTK) | directly druggable gene/pathway |  |  |
| IGF2 | Receptor Tyrosine Kinase (RTK) | indirectly actionable gene (IGF1R) |  | metformin, ganitumab |
| IGF1R | Receptor Tyrosine Kinase (RTK) | directly druggable gene/gene family | ceritinib | ganitumab, linsitinib |
| IGF1 | Receptor Tyrosine Kinase (RTK) | indirectly actionable gene (IGF1R) |  | ganitumab, linsitinib |
| PTCH1 | Sonic Hedgehog (SHH) | indirectly actionable gene (SMO) |  | vismodegib |
| GLI1 | Sonic Hedgehog (SHH) | indirectly actionable gene (SMO) |  | vismodegib |
| SMO | Sonic Hedgehog (SHH) | directly druggable gene/pathway | vismodegib, sonidegib |  |
| CDK9 | Transcription | directly druggable gene/gene family |  | dinaciclib |
| VEGFA | VEGFR | directly druggable gene/gene family | bevacizumab, pegaptanib, ranibizumab, ziv-aflibercept, lenvatinib |  |
| VEGFC | VEGFR | indirectly actionable gene (KDR) |  | pazopanib |
| WEE1 | WEE1 | directly druggable gene/gene family |  | adavosertib |

#### **Table A3. CARE IMPACT findings by sample**

| **Patient ID** | **Sample ID** | **CARE IMPACT Finding** | **Tumor Vulnerability Category** | **Therapies** | **Clinician Prioritization** | **Prioritized Finding Outcome** | **CARE IMPACT Finding Source** | **Pan Disease Outlier Source** | **Pan Disease Cohorts** | **Notes** | **Compendium Version** |
| --- | --- | --- | --- | --- | --- | --- | --- | --- | --- | --- | --- |
| TH34_1239 | TH34_1239_S01 | PIK3CD | PI3K/AKT/mTOR | Everolimus | Accepted, Deferred, Received therapy with more published evidence of efficacy |  | pan-cancer outlier |  |  |  | compendium-v7 |
| TH34_1239 | TH34_1239_S01 | NRAS | RAS/RAF/MEK | Trametinib or Selumetinib | Accepted, Deferred, Received therapy with more published evidence of efficacy |  | expressed mutation |  |  |  | compendium-v7 |
| TH34_1238 | TH34_1238_S01 | mTOR | PI3K/AKT/mTOR | Everolimus | Accepted, Deferred, Received therapy with more published evidence of efficacy |  | pan-cancer outlier |  |  |  | compendium-v7 |
| TH34_1238 | TH34_1238_S01 | KRAS | RAS/RAF/MEK | Trametinib or Selumetinib | Accepted, Deferred, Received therapy with more published evidence of efficacy |  | expressed mutation |  |  |  | compendium-v7 |
| TH34_1350 | TH34_1350_S01 | PIK3R5 | PI3K/AKT/mTOR | Everolimus | Accepted, Deferred, Received therapy with more published evidence of efficacy |  | pan-cancer outlier |  |  |  | compendium-v8 |
| TH34_1350 | TH34_1350_S01 | MEK | RAS/RAF/MEK | Trametinib or Selumetinib | Accepted, Deferred, Received therapy with more published evidence of efficacy |  | other highly expressed gene |  |  |  | compendium-v8 |
| TH34_1381 | TH34_1381_S01 | VEGFA | VEGF | Bevacizumab | Accepted, Deferred, Received therapy with more published evidence of efficacy |  | pan-cancer outlier, pan-disease outlier | consensus | canonical |  | compendium-v8 |
| TH34_1381 | TH34_1381_S01 | ERBB2 | ERBB | Lapatinib | Accepted, Prioritized, Implemented | Rapid disease progression | pan-disease outlier | consensus | canonical |  | compendium-v8 |
| TH34_1381 | TH34_1381_S01 | PARP1 | PARP | Olaparib | Declined, Not FDA approved |  | pan-disease outlier | consensus | canonical |  | compendium-v8 |
| TH34_2410 | TH34_2410_S01 | MAPK11 | p38 | Nilotinib | Accepted, Deferred, Standard of care |  | pan-disease outlier | consensus | curated | curated cohorts used: uterine cohort (uterine corpus endometrioid carcinoma, uterine carcinosarcoma), soft tissue sarcoma; outlier identified by original canonical analysis with as a pd outlier but upon reanalysis with curated sarcoma cohort it was not identified as an outlier; the reason a reanalysis was done after initially presenting to clinicians was that the clinicians thought this sample was more similar to sarcomas as opposed to uterine carcinomas | compendium-v11_polya |
| TH34_2410 | TH34_2410_S01 | ESR1 | Estrogen Receptor | Palbociclib | Accepted, Deferred, Standard of care |  | pan-cancer outlier |  |  | curated cohorts used: uterine cohort (uterine corpus endometrioid carcinoma, uterine carcinosarcoma), soft tissue sarcoma; outlier identified by original canonical analysis with as a pc outlier but upon reanalysis with curated sarcoma cohort it was identified as both pc and pd; the reason a reanalysis was done after initially presenting to clinicians was that the clinicians thought this sample was more similar to sarcomas as opposed to uterine carcinomas | compendium-v11_polya |
| TH34_2410 | TH34_2410_S01 | ABCB1 | ABCB1 | Regorafenib or Ulixertinib | Accepted, Deferred, Standard of care |  | pan-disease outlier | consensus | curated | curated cohorts used: uterine cohort (uterine corpus endometrioid carcinoma, uterine carcinosarcoma), soft tissue sarcoma; outlier identified by original canonical analysis with as a pc outlier but upon reanalysis with curated sarcoma cohort it was identified as both pc and pd; the reason a reanalysis was done after initially presenting to clinicians was that the clinicians thought this sample was more similar to sarcomas as opposed to uterine carcinomas | compendium-v11_polya |
| TH34_1162 | TH34_1162_S01 | CSF1R | RTK | Pazopanib | Accepted, Deferred, Received therapy with more published evidence of efficacy |  | pan-disease outlier | consensus | canonical |  | compendium-v7 |
| TH34_1162 | TH34_1162_S01 | JAK1 | JAK/STAT | Ruxolitinib | Accepted, Deferred, Received therapy with more published evidence of efficacy |  | pan-cancer outlier |  |  |  | compendium-v7 |
| TH34_2411 | TH34_2411_S01 | MDM2 | MDM2/MDMX | ALRN-6924 | Declined, Not FDA approved |  | pan-cancer outlier, pan-disease outlier | consensus | canonical |  | compendium-v11_polya |
| TH34_2411 | TH34_2411_S01 | B4GALNT1 | B4GALNT1 | Dinutuximab | Accepted, Deferred, Received therapy with more published evidence of efficacy |  | pan-cancer outlier, pan-disease outlier | consensus | canonical |  | compendium-v11_polya |
| TH34_2411 | TH34_2411_S01 | CDK4 | Cell Cycle | Palbociclib | Accepted, Deferred, Received therapy with more published evidence of efficacy |  | pan-cancer outlier, pan-disease outlier | consensus | canonical |  | compendium-v11_polya |
| TH34_2411 | TH34_2411_S01 | MAP2K2 | RAS/RAF/MEK | Trametinib or Selumetinib | Accepted, Deferred, Received therapy with more published evidence of efficacy |  | pan-cancer outlier, pan-disease outlier | consensus | canonical |  | compendium-v11_polya |
| TH34_2666 | TH34_2666_S01 | MDM2 | MDM2/MDMX | APG-115 or ALRN-6924 | Declined, Not FDA approved |  | pan-cancer outlier, pan-disease outlier | single cohort | curated | curated cohort used: soft tissue sarcoma | compendium-v11_polya |
| TH34_2666 | TH34_2666_S01 | FLT4 | RTK | Pazopanib | Accepted, Deferred, No further treatment needed |  | pan-cancer outlier, pan-disease outlier | single cohort | curated | curated cohort used: soft tissue sarcoma | compendium-v11_polya |
| TH34_1163 | TH34_1163_S01 | IGF2 | IGF1R | Metformin or Ganitumab | Declined, Evidence against Treehouse therapy |  | pan-cancer outlier |  |  |  | compendium-v7 |
| TH34_1163 | TH34_1163_S01 | HIF1 signaling/HMOX1 | Other: HMOX1/HIF1A Hypoxia Signaling | Nitroglycerin or EZN-2208 | Accepted, Deferred, Received therapy with more published evidence of efficacy |  | pan-disease outlier | consensus | canonical |  | compendium-v7 |
| TH34_1149 | TH34_1149_S02 | NTRK3 | NTRK | Entrectinib | Declined, Evidence against Treehouse therapy |  | pan-cancer outlier |  |  |  | compendium-v7 |
| TH34_1150 | TH34_1150_S02 | CDK9 | CDK | Dinaciclib | Declined, Not FDA approved |  | pan-disease outlier | consensus | canonical |  | compendium-v8 |
| TH34_1150 | TH34_1150_S02 | PDCD1 | T-Cell | Pembrolizumab | Declined, Evidence against Treehouse therapy |  | pan-cancer outlier |  |  |  | compendium-v8 |
| TH34_1150 | TH34_1150_S02 | JAK1 | JAK/STAT | Ruxolitinib | Accepted, Deferred, Received therapy with more published evidence of efficacy |  | pan-cancer outlier |  |  |  | compendium-v8 |
| TH34_1179 | TH34_1179_S01 | FGFR1 | RTK | Pazopanib | Accepted, Deferred, Standard of care |  | pan-cancer outlier |  |  |  | compendium-v7 |
| TH34_1179 | TH34_1179_S01 | KRAS | RAS/RAF/MEK | Trametinib or Selumetinib | Accepted, Deferred, Standard of care |  | expressed mutation |  |  |  | compendium-v7 |
| TH34_1179 | TH34_1179_S01 | Hedgehog signaling | Other: Hedgehog signaling | Vismodegib | Accepted, Deferred, Standard of care |  | pan-cancer outlier |  |  |  | compendium-v7 |
| TH34_1240 | TH34_1240_S01 | EWSR1-FLI1 | EWSR1-FLI1 | MYC-targeted drugs (clinical trials for INCB05987 (NCT0351440) or TK216 (NCT02657005)) | Accepted, Prioritized, Not implemented | Rapid disease progression | expressed fusion |  |  |  | compendium-v7 |
| TH34_1349 | TH34_1349_S01 | KIT | KIT | Imatinib or Sunitinib or Regorafenib | Accepted, Prioritized, Implemented | Response | pan-cancer outlier, pan-disease outlier | single cohort | curated | curated cohorts used: soft tissue sarcoma & stomach | compendium-v8 |
| TH34_1349 | TH34_1349_S01 | IGF2, IGF1R | IGF1R | Linsitinib | Declined, Not FDA approved |  | pan-cancer outlier |  |  | curated cohorts used: soft tissue sarcoma & stomach | compendium-v8 |
| TH34_1349 | TH34_1349_S01 | ETV1 | RAS/RAF/MEK | Trametinib or Selumetinib | Accepted, Deferred, Received therapy with more published evidence of efficacy |  | pan-cancer outlier, pan-disease outlier | single cohort | curated | curated cohorts used: soft tissue sarcoma & stomach | compendium-v8 |
| TH34_1349 | TH34_1349_S02 | KIT | KIT | Imatinib or Sunitinib or Regorafenib | Accepted, Prioritized, Implemented | Response | pan-cancer outlier, pan-disease outlier | single cohort | curated | curated cohorts used: soft tissue sarcoma & stomach | compendium-v8 |
| TH34_1349 | TH34_1349_S02 | ETV1 | RAS/RAF/MEK | Trametinib or Selumetinib | Accepted, Deferred, Received therapy with more published evidence of efficacy |  | pan-cancer outlier, pan-disease outlier | single cohort | curated | curated cohorts used: soft tissue sarcoma & stomach | compendium-v8 |
| TH34_1349 | TH34_1349_S02 | PTCH1 | Hedgehog Signaling | Vismodegib | Accepted, Deferred, Received therapy with more published evidence of efficacy |  | pan-cancer outlier, pan-disease outlier | single cohort | curated | curated cohorts used: soft tissue sarcoma & stomach | compendium-v8 |
| TH34_1351 | TH34_1351_S01 | MEK | RAS/RAF/MEK | Trametinib or Selumetinib | Accepted, Deferred, Received therapy with more published evidence of efficacy |  | other highly expressed gene |  |  |  | compendium-v8 |
| TH34_1351 | TH34_1351_S01 | MAP2K4 | JNK | Vemurafenib | Accepted, Deferred, Received therapy with more published evidence of efficacy |  | pan-cancer outlier, pan-disease outlier | consensus | canonical |  | compendium-v8 |
| TH34_1352 | TH34_1352_S01 | FGFR1, FGFR2, PDGFRA | RTK | Pazopanib | Accepted, Prioritized, Implemented | Progression | pan-cancer outlier, pan-disease outlier | single cohort | canonical |  | compendium-v8 |
| TH34_1352 | TH34_1352_S01 | CCND2 | Cell Cycle | Ribociclib | Accepted, Prioritized, Implemented | Response | pan-disease outlier | single cohort | canonical |  | compendium-v8 |
| TH34_1379 | TH34_1379_S01 | IGF2, IGF1R | IGF1R | Ganitumab | Declined, Not FDA approved |  | pan-cancer outlier, pan-disease outlier | consensus | curated | curated cohorts used: liver and small round blue cell; outlier identified by both curated cohorts and one canonical cohort | compendium-v8 |
| TH34_1379 | TH34_1379_S01 | FGFR1 | RTK | Pazopanib | Accepted, Prioritized, Not implemented | Rapid disease progression | pan-disease outlier | consensus | curated | curated cohorts used: liver and small round blue cell; outlier identified by both curated cohorts and one canonical cohort | compendium-v8 |
| TH34_1379 | TH34_1379_S01 | CCND2, CDK4 | Cell Cycle | Ribociclib | Accepted, Deferred, Received therapy with more published evidence of efficacy |  | pan-disease outlier | consensus | curated | curated cohorts used: liver and small round blue cell; outlier not identified by small round blue cell cohort | compendium-v8 |
| TH34_1380 | TH34_1380_S01 | IGF2, IGF1R | IGF1R | Ganitumab | Declined, Not FDA approved |  | pan-cancer outlier |  |  |  | compendium-v8 |
| TH34_1380 | TH34_1380_S01 | NOTCH3 | NOTCH | Tarextumab | Accepted, Prioritized, Not implemented | Waiting for treatment approval | pan-cancer outlier, pan-disease outlier | consensus | canonical |  | compendium-v8 |
| TH34_1399 | TH34_1399_S01 | VEGFA | VEGF | Bevacizumab | Accepted, Deferred, Received therapy with more published evidence of efficacy |  | pan-disease outlier | consensus | curated | curated cohorts used: soft tissue sarcoma | compendium-v8 |
| TH34_1399 | TH34_1399_S01 | IGF1, IGF2 | IGF1R | Ganitumab | Declined, Not FDA approved |  | pan-cancer outlier |  |  |  | compendium-v8 |
| TH34_1400 | TH34_1400_S01 | STAT5A | JAK/STAT | Ruxolitinib | Accepted, Deferred, No further treatment needed |  | pan-disease outlier | consensus | curated | curated cohort used: dedifferentiated liposarcoma | compendium-v8 |
| TH34_1412 | TH34_1412_S01 | NTRK3 | NTRK | Entrectinib | Declined, Evidence against Treehouse therapy |  | pan-cancer outlier |  |  | curated cohort used: soft tissue sarcoma | compendium-v8 |
| TH34_1412 | TH34_1412_S01 | IGF2 | IGF1R | Ganitumab | Declined, Not FDA approved |  | pan-cancer outlier |  |  | curated cohort used: soft tissue sarcoma | compendium-v8 |
| TH34_1412 | TH34_1412_S01 | KDR | RTK | Pazopanib | Declined, Evidence against Treehouse therapy |  | pan-cancer outlier, pan-disease outlier | single cohort | curated | curated cohort used: soft tissue sarcoma | compendium-v8 |
| TH34_1412 | TH34_1412_S01 | ERBB2 | ERBB | Trastuzumab | Accepted, Deferred, Received therapy with more published evidence of efficacy |  | pan-disease outlier | single cohort | curated | curated cohort used: soft tissue sarcoma | compendium-v8 |
| TH34_1414 | TH34_1414_S01 | ALK | ALK | Crizotinib | Accepted, Deferred, Received therapy with more published evidence of efficacy |  | pan-cancer outlier |  |  |  | compendium-v8 |
| TH34_1414 | TH34_1414_S01 | IGF2 | IGF1R | Ganitumab | Declined, Not FDA approved |  | pan-cancer outlier |  |  |  | compendium-v8 |
| TH34_1414 | TH34_1414_S01 | FGFR4 | RTK | Ponatinib | Accepted, Deferred, Received therapy with more published evidence of efficacy |  | pan-cancer outlier |  |  |  | compendium-v8 |
| TH34_1414 | TH34_1414_S01 | CDK4 | Cell Cycle | Ribociclib | Accepted, Deferred, Received therapy with more published evidence of efficacy |  | pan-cancer outlier |  |  |  | compendium-v8 |
| TH34_1415 | TH34_1415_S01 | FLT4 | RTK | Pazopanib | Accepted, Deferred, Received therapy with more published evidence of efficacy |  | pan-cancer outlier, pan-disease outlier | consensus | curated | curated cohort used: soft tissue sarcoma | compendium-v8 |
| TH34_1415 | TH34_1415_S01 | IL6 | IL6 | Siltuximab | Accepted, Deferred, Received therapy with more published evidence of efficacy |  | pan-cancer outlier |  |  |  | compendium-v8 |
| TH34_1444 | TH34_1444_S01 | AKT1, AKT2 | PI3K/AKT/mTOR | Everolimus | Accepted, Deferred, Standard of care |  | pan-cancer outlier, pan-disease outlier | consensus | canonical |  | compendium-v8 |
| TH34_1444 | TH34_1444_S01 | VEGFC | RTK (VEGFR) | Pazopanib | Accepted, Deferred, Standard of care |  | pan-cancer outlier, pan-disease outlier | consensus | canonical |  | compendium-v8 |
| TH34_1444 | TH34_1444_S01 | KIT | KIT | Sunitinib | Accepted, Deferred, Standard of care |  | pan-cancer outlier, pan-disease outlier | consensus | canonical |  | compendium-v8 |
| TH34_1445 | TH34_1445_S02 | WEE1 | WEE1 | Adavosertib | Accepted, Deferred, Received therapy with more published evidence of efficacy |  | pan-cancer outlier |  |  |  | compendium-v9 |
| TH34_1445 | TH34_1445_S02 | VEGFA | VEGF | Bevacizumab | Accepted, Deferred, Received therapy with more published evidence of efficacy |  | pan-disease outlier | consensus | curated | curated cohort used: glioma | compendium-v9 |
| TH34_1445 | TH34_1445_S02 | NTRK3 | NTRK | Entrectinib | Declined, Evidence against Treehouse therapy |  | pan-cancer outlier |  |  |  | compendium-v9 |
| TH34_1445 | TH34_1445_S02 | IGF2 | IGF1R | Ganitumab | Declined, Not FDA approved |  | pan-cancer outlier, pan-disease outlier | consensus | curated |  | compendium-v9 |
| TH34_1446 | TH34_1446_S01 | NTRK3 | NTRK | Entrectinib | Accepted, Deferred, Received therapy with more published evidence of efficacy |  | pan-cancer outlier |  |  |  | compendium-v9 |
| TH34_1447 | TH34_1447_S01 | PSMB5, PSMC1, PSMD4 | Proteasome | Bortezomib | Accepted, Deferred, Received therapy with more published evidence of efficacy |  | pan-cancer outlier, pan-disease outlier | consensus | curated | curated cohort used: soft tissue sarcoma; outlier was identified by curated cohort and presented to clinicians, however, there was discussion among the Treehouse team about whether or not to present it as a finding | compendium-v9 |
| TH34_1447 | TH34_1447_S01 | CCNE1 | Cell Cycle | Dinaciclib | Declined, Not FDA approved |  | pan-disease outlier | consensus | curated |  | compendium-v9 |
| TH34_1447 | TH34_1447_S01 | PARP1, PARP2 | PARP | Olaparib | Declined, Not FDA approved |  | pan-cancer outlier, pan-disease outlier | consensus | curated | curated cohort used: soft tissue sarcoma | compendium-v9 |
| TH34_1447 | TH34_1447_S02 | PARP2 | PARP | Olaparib | Declined, Not FDA approved |  | pan-cancer outlier |  |  |  | compendium-v9 |
| TH34_1447 | TH34_1447_S02 | PSMC1, PSMD4 | Proteasome | Bortezomib | Accepted, Deferred, Received therapy with more published evidence of efficacy |  | pan-cancer outlier, pan-disease outlier | single cohort | curated | curated cohort used: soft tissue sarcoma; outlier was identified by curated cohort and presented to clinicians, however, there was discussion among the Treehouse team about whether or not to present it as a finding | compendium-v9 |
| TH34_1447 | TH34_1447_S02 | IGF2 | IGF1R | Ganitumab | Declined, Not FDA approved |  | pan-cancer outlier |  |  |  | compendium-v9 |
| TH34_1452 | TH34_1452_S01 | PIK3R2 | PI3K/AKT/mTOR | Everolimus | Accepted, Deferred, Received therapy with more published evidence of efficacy |  | pan-cancer outlier |  |  |  | compendium-v9 |
| TH34_1452 | TH34_1452_S01 | IGF2 | IGF1R | Ganitumab | Declined, Not FDA approved |  | pan-cancer outlier |  |  |  | compendium-v9 |
| TH34_1452 | TH34_1452_S01 | FGFR3 | RTK | Pazopanib | Accepted, Deferred, Received therapy with more published evidence of efficacy |  | pan-cancer outlier |  |  |  | compendium-v9 |
| TH34_1452 | TH34_1452_S01 | HDAC4 | HDAC | Vorinostat | Accepted, Deferred, Received therapy with more published evidence of efficacy |  | pan-cancer outlier, pan-disease outlier | consensus | canonical |  | compendium-v9 |
| TH34_1455 | TH34_1455_S01 | CDK9 | CDK | Dinaciclib | Declined, Not FDA approved |  | pan-cancer outlier |  |  |  | compendium-v9 |
| TH34_1455 | TH34_1455_S01 | IGF2 | IGF1R | Ganitumab | Declined, Not FDA approved |  | pan-cancer outlier, pan-disease outlier | consensus | canonical |  | compendium-v9 |
| TH34_1455 | TH34_1455_S01 | ERBB2 | ERBB | Neratinib | Accepted, Prioritized, Not implemented | Therapy unavailable | pan-disease outlier | consensus | canonical |  | compendium-v9 |
| TH34_1455 | TH34_1455_S01 | FGFR3 | RTK | Pazopanib | Accepted, Deferred, Received therapy with more published evidence of efficacy |  | pan-disease outlier | consensus | canonical |  | compendium-v9 |
| TH34_1456 | TH34_1456_S02 | GATA2 | Proteasome | Bortezomib | Accepted, Deferred, Received therapy with more published evidence of efficacy |  | pan-cancer outlier, pan-disease outlier | consensus | canonical |  | compendium-v9 |
| TH34_1456 | TH34_1456_S02 | KDR | RTK | Pazopanib | Accepted, Prioritized, Implemented | Response | pan-cancer outlier, pan-disease outlier | consensus | canonical |  | compendium-v9 |
| TH34_1456 | TH34_1456_S02 | HDAC7 | HDAC | Vorinostat | Accepted, Deferred, Received therapy with more published evidence of efficacy |  | pan-disease outlier | consensus | canonical |  | compendium-v9 |
| TH34_2292 | TH34_2292_S01 | VEGFA | VEGF | Bevacizumab | Accepted, Prioritized, Not implemented | Rapid disease progression | pan-cancer outlier |  |  |  | compendium-v10_polya |
| TH34_2292 | TH34_2292_S01 | FLT4 | RTK | Pazopanib | Accepted, Deferred, Unable to take oral medication |  | pan-cancer outlier |  |  |  | compendium-v10_polya |
| TH34_2293 | TH34_2293_S01 | CDK1 | CDK1 | Dinaciclib | Declined, Not FDA approved |  | other highly expressed gene |  |  |  | compendium-v10_polya |
| TH34_2293 | TH34_2293_S01 | NOTCH3 | NOTCH | Nirogacestat | Declined, Not FDA approved |  | other highly expressed gene |  |  |  | compendium-v10_polya |
| TH34_2351 | TH34_2351_S01 | HIF1 signaling/HMOX1 | Other: HMOX1/HIF1A Hypoxia Signaling | Nitroglycerin | Declined, No phase I data in children |  | pan-disease outlier | consensus | canonical |  | compendium-v10_polya |
| TH34_2351 | TH34_2351_S01 | MAP2K1 | MAPK | Trametinib or Selumetinib | Accepted, Prioritized, Implemented | Progression | other highly expressed gene |  |  |  | compendium-v10_polya |

**Table A4. DNA variant calls per patient sample**

| **Sample ID** | **Gene** | **Mutation** | **Note** | **Test vendor** | **FDA approved therapies in patients tumor type** | **FDA approved therapies in another tumor type** | **Potential clinical trials** | **Potentially actionable** | **Indication status** |
| --- | --- | --- | --- | --- | --- | --- | --- | --- | --- |
| TH34_1150_S02 | EWSR1 | EWSR1-FLI1 fusion (type 1) |  | Foundation medicine | No | No | Yes | Yes |  |
| TH34_1162_S01 | EWSR1 | EWSR1-FLI1 fusion (type 3) |  | Foundation medicine | No | No | Yes | Yes |  |
| TH34_1163_S01 | FBXW7 | E471G |  | Foundation medicine | No | Temsirolimus, Everolimus | Yes | Yes | indication |
| TH34_1163_S01 | FBXW7 | R465H |  | Foundation medicine | No | Temsirolimus, Everolimus | Yes | Yes | indication |
| TH34_1163_S01 | KRAS | amplification |  | Foundation medicine | No | Cobimetinib, Trametinib, Binimetinib | Yes | Yes | indication |
| TH34_1163_S01 | KRAS | Q61P |  | Foundation medicine | No | Cobimetinib, Trametinib, Binimetinib | Yes | Yes | indication |
| TH34_1163_S01 | CDKN2A | loss |  | Foundation medicine | No | No | No | No |  |
| TH34_1163_S01 | CDKN2B | loss |  | Foundation medicine | No | No | No | No |  |
| TH34_1163_S01 | TP53 | R175fs*7 |  | Foundation medicine | No | No | No | No |  |
| TH34_1179_S01 | KRAS | G12D |  | Foundation medicine | Cetuximab, Panitumumab | No | Yes | Yes | counter indication |
| TH34_1179_S01 | PIK3CA | E545K | subclonal | Foundation medicine | No | No | Yes | Yes |  |
| TH34_1179_S01 | PTEN | loss |  | Foundation medicine | No | No | Yes | Yes |  |
| TH34_1179_S01 | APC | S1465fs*3 |  | Foundation medicine | No | No | No | No |  |
| TH34_1179_S01 | APC | T829fs*13 |  | Foundation medicine | No | No | No | No |  |
| TH34_1179_S01 | KDM6A | R1213* |  | Foundation medicine | No | No | No | No |  |
| TH34_1179_S01 | TERT | -124C>T | promoter | Foundation medicine | No | No | No | No |  |
| TH34_1240_S01 | EWSR1 | EWSR1-FLI1 fusion (type 1) |  | Foundation medicine | No | No | Yes | Yes |  |
| TH34_1240_S01 | MYC | amplification |  | Foundation medicine | No | No | Yes | Yes |  |
| TH34_1240_S01 | TP53 | R248Q |  | Foundation medicine | No | No | No | No |  |
| TH34_1349_S01 | None found |  | mutation testing was done on second sample from patient (TH34_1349_S02) | STAMP |  |  |  |  |  |
| TH34_1349_S02 | None found |  | no clinically relevant variants reported | STAMP |  |  |  |  |  |
| TH34_1350_S01 | CBL | Y371H |  | Foundation medicine | No | No | No | No |  |
| TH34_1351_S01 | C17orf39 | amplification | equivocal | Foundation medicine | No | No | No | No |  |
| TH34_1351_S01 | MSH3 | 580-2A>G | splice site | Foundation medicine | No | No | No | No |  |
| TH34_1351_S01 | STAG2 | P456A |  | Foundation medicine | No | No | No | No |  |
| TH34_1352_S01 | SMARCB1 | loss |  | Foundation medicine | No | No | Yes | Yes |  |
| TH34_1379_S01 | CTNNB1 | V22_A39del |  | Foundation medicine | No | No | Yes | Yes |  |
| TH34_1380_S01 | CCND1 | amplification | equivocal | Foundation medicine | No | Abemaciclib, Palbociclib, Ribociclib | Yes | Yes | indication |
| TH34_1380_S01 | FGF19 | amplification | equivocal | Foundation medicine | No | No | No | No |  |
| TH34_1380_S01 | FGF3 | amplification | equivocal | Foundation medicine | No | No | No | No |  |
| TH34_1380_S01 | FGF4 | amplification | equivocal | Foundation medicine | No | No | No | No |  |
| TH34_1380_S01 | TP63 | P464fs*3 |  | Foundation medicine | No | No | No | No |  |
| TH34_1381_S01 | IKBKE | amplification | equivocal | Foundation medicine | No | No | No | No |  |
| TH34_1381_S01 | MCL1 | amplification | equivocal | Foundation medicine | No | No | No | No |  |
| TH34_1381_S01 | NTRK1 | amplification | equivocal | Foundation medicine | No | No | No | No |  |
| TH34_1399_S01 | EWSR1 | EWSR1-WT1 fusion |  | Foundation medicine | No | No | Yes | Yes |  |
| TH34_1399_S01 | LRP1B | R2234fs*30 |  | Foundation medicine | No | No | No | No |  |
| TH34_1412_S01 | EWSR1 | EWSR1-WT1 fusion |  | Foundation medicine | No | No | Yes | Yes |  |
| TH34_1414_S01 | CDK4 | amplification |  | Foundation medicine | No | Palbociclib, Ribociclib | Yes | Yes | indication |
| TH34_1414_S01 | FGF14 | amplification |  | Foundation medicine | No | No | No | No |  |
| TH34_1414_S01 | NOTCH1 | splice site 6181-1G>A |  | Foundation medicine | No | No | No | No |  |
| TH34_1414_S01 | IRS2 | amplification |  | Foundation medicine | No | No | No | No |  |
| TH34_1414_S01 | PAX3 | PAX3-FOXO1 fusion |  | Foundation medicine | No | No | No | No |  |
| TH34_1415_S01 | ARID1A | ARID1A-PIGV rearrangement |  | Foundation medicine | No | No | No | No |  |
| TH34_1415_S01 | CIC | CIC-DUX4 fusion |  | Foundation medicine | No | No | No | No |  |
| TH34_1415_S01 | EBF1 | V445M |  | Foundation medicine | No | No | No | No |  |
| TH34_1444_S01 | RB1 | L486fs*9 |  | Foundation medicine | No | No | No | No |  |
| TH34_1445_S02 | H3F3A | K28M |  | Foundation medicine | No | No | No | No |  |
| TH34_1445_S02 | PIK3C2B | amplification | equivocal | Foundation medicine | No | No | No | No |  |
| TH34_1445_S02 | BRAF | V600E |  | Foundation medicine | No | Binimetinib, Cobimetinib, Dabrafenib, Encorafenib, Regorafenib, Trametinib, Vemurafenib | Yes | Yes | indication |
| TH34_1446_S01 | BRAF | V600E |  | Foundation medicine | No | Binimetinib, Cobimetinib, Dabrafenib, Encorafenib, Regorafenib, Trametinib, Vemurafenib | Yes | Yes | indication |
| TH34_1447_S01 | PDGFRB | N666K | mutation testing was done on a metastatic lung sample | Foundation medicine | No | Sunitinib | Yes | Yes | indication |
| TH34_1447_S01 | LRRK2 | R1639fs*15 | mutation testing was done on a metastatic lung sample | Foundation medicine | No | No | No | No |  |
| TH34_1447_S01 | NFKBIA | amplification | mutation testing was done on a metastatic lung sample | Foundation medicine | No | No | No | No |  |
| TH34_1447_S01 | NKX2-1 | amplification | mutation testing was done on a metastatic lung sample | Foundation medicine | No | No | No | No |  |
| TH34_1447_S01 | SDHA | splice site 150+1G>A | mutation testing was done on a metastatic lung sample | Foundation medicine | No | No | No | No |  |
| TH34_1447_S01 | TP53 | C238S | mutation testing was done on a metastatic lung sample | Foundation medicine | No | No | No | No |  |
| TH34_1447_S02 | PDGFRB | N666K | mutation testing was done on a metastatic lung sample | Foundation medicine | No | Sunitinib | Yes | Yes | indication |
| TH34_1447_S02 | LRRK2 | R1639fs*15 | mutation testing was done on a metastatic lung sample | Foundation medicine | No | No | No | No |  |
| TH34_1447_S02 | NFKBIA | amplification | mutation testing was done on a metastatic lung sample | Foundation medicine | No | No | No | No |  |
| TH34_1447_S02 | NKX2-1 | amplification | mutation testing was done on a metastatic lung sample | Foundation medicine | No | No | No | No |  |
| TH34_1447_S02 | SDHA | splice site 150+1G>A | mutation testing was done on a metastatic lung sample | Foundation medicine | No | No | No | No |  |
| TH34_1447_S02 | TP53 | C238S | mutation testing was done on a metastatic lung sample | Foundation medicine | No | No | No | No |  |
| TH34_1452_S01 | None found |  | no clinically relevant variants reported | Foundation medicine |  |  |  |  |  |
| TH34_1455_S01 | None found |  | no clinically relevant variants reported | Foundation medicine |  |  |  |  |  |
| TH34_1456_S02 | BIRC3 | amplification |  | Foundation medicine | No | No | No | No |  |
| TH34_1456_S02 | CCND3 | amplification |  | Foundation medicine | No | No | No | No |  |
| TH34_1456_S02 | TP53 | loss |  | Foundation medicine | No | No | No | No |  |
| TH34_2292_S01 | CIC | rearrangement | exon 20 | Foundation medicine | No | No | No | No |  |
| TH34_2293_S01 | EWSR1 | EWSR1-CREB3L1 fusion |  | Foundation medicine | No | No | No | No |  |
| TH34_2293_S01 | NOTCH1 | splice site 5167+1G>A | subclonal | Foundation medicine | No | No | No | No |  |
| TH34_2293_S01 | TMEM30A | Y134fs*1 |  | Foundation medicine | No | No | No | No |  |
| TH34_2410_S01 | CDKN2A | I49T |  | Foundation medicine | No | No | No | No |  |
| TH34_2411_S01 | ATRX | deletion exon 23 |  | Foundation medicine | No | No | No | No |  |
| TH34_2411_S01 | FRS2 | amplification |  | Foundation medicine | No | No | No | No |  |
| TH34_2411_S01 | MCL1 | L21_G24del |  | Foundation medicine | No | No | No | No |  |
| TH34_2411_S01 | CDK4 | amplification |  | Foundation medicine | No | Palbociclib, Ribociclib | Yes | Yes | indication |
| TH34_2411_S01 | MDM2 | amplification |  | Foundation medicine | No | No | Yes | Yes |  |
| TH34_2666_S01 | CIC | CIC-DUX4 fusion |  | Foundation medicine | No | No | No | No |  |
| **No mutation report** | | | | | | | | | |
| TH34_1149_S02 |  |  | sample failed testing process; low sample quality | Foundation medicine |  |  |  |  |  |
| TH34_1400_S01 |  |  | testing not done; low sample quantity |  |  |  |  |  |  |
| TH34_1239_S01 |  |  | testing not done; low sample quantity |  |  |  |  |  |  |
| TH34_1238_S01 |  |  | testing not done |  |  |  |  |  |  |
| TH34_2351_S01 |  |  | testing not done; not covered by patient’s insurance |  |  |  |  |  |  |

#### **Table A5. Published repository datasets included in the Treehouse Compendia**

|  |  |  |  |  | **Number of Samples (N) per PolyA Compendium Version** | | | | |
| --- | --- | --- | --- | --- | --- | --- | --- | --- | --- |
| **TH Dataset** | **Accession/Partner** | **Project** | **Repository/ Partner** | **Disease** | **v7**  May 2018 | **v8**  Jul 2018 | **v9**  Mar 2019 | **v10**  Jul 2019 | **v11**  Apr 2020 |
| TARGET | phs000218 | TARGET | dbGap | multiple cancers | 784 | 784 | 784 | 784 | 1190 |
| TCGA | phs000178 | TCGA | dbGap | multiple cancers | 9806 | 9806 | 9806 | 9806 | 9806 |
| THR08 | EGAD00001001098 | St Jude PCGP | EGA | ALL with MLL rearrangements | 63 | 63 | 63 | 63 | 63 |
| THR09 | EGAD00001000356 | ICGC | EGA | B-cell lymphoma | 23 | 23 | 23 | 23 | 23 |
| THR10 | EGAD00001002680 | St Jude PCGP | EGA | adrenocortical carcinoma | 15 | 15 | 15 | 15 | 15 |
| THR11 | EGAD00001001666 | St Jude PCGP | EGA | low grade glioma | 25 | 25 | 25 | 25 | 25 |
| THR12 | phs000709.v1.p1 | n/a | dbGap | fibrolamellar hepatocellular carcinoma | 0 | 0 | 20 | 0 | 0 |
| THR13 | phs000900.v1.p1 | PBTC | dbGap | DIPG | 10 | 10 | 10 | 10 | 10 |
| THR14 | CBTTC | CBTTC | Cavatica | high grade glioma | 29 | 81 | 29 | 29 | 29 |
| THR15 | phs000699.v1.p1 | n/a | dbGap | osteosarcoma | 29 | 29 | 29 | 29 | 29 |
| THR17 | EGAD00001000158 | ICGC/MAGIC | EGA | medulloblastoma | 17 | 17 | 17 | 17 | 17 |
| THR18 | EGAD00001000328 | ICGC | EGA | medulloblastoma | 6 | 6 | 6 | 6 | 6 |
| THR19 | EGAD00001000617 | ICGC | EGA | pilocytic astrocytoma | 23 | 23 | 23 | 23 | 23 |
| THR20 | EGAD00001001620 | ICGC | EGA | medulloblastoma | 39 | 39 | 39 | 39 | 39 |
| THR21 | EGAD00001001944 | ICGC | EGA | pediatric glioblastoma | 0 | 0 | 41 | 0 | 0 |
| THR22 | EGAD00001000648 | ICGC | EGA | germinal B-cell lymphoma | 24 | 24 | 24 | 24 | 24 |
| THR24 | SJC-DS-1001 | St Jude PCGP | EGA | multiple cancers | 0 | 0 | 0 | 724 | 709 |
| THR25 | EGAD00001000826 | ICGC | EGA | osteosarcoma | 7 | 7 | 7 | 7 | 7 |
| THR28 | SRP040454 | Kohsaka et al | SRA | rhabdomyosarcoma | 3 | 3 | 3 | 3 | 3 |
| THR29 | phs000720.v2.p1 | Shern et al | dbGap | rhabdomyosarcoma | 84 | 84 | 84 | 84 | 84 |
| THR30 | phs000768.v2.p1 | Brohl et al | dbGap | Ewing sarcoma | 62 | 62 | 62 | 62 | 62 |
| THR31 | phs000673.v2.p1 | Peds Mi-OncoSeq/Met500 | dbGap | multiple cancers | 75 | 75 | 87 | 87 | 87 |
| THR32 | EGAD00001001927 | ICGC | EGA | CNS-PNETs | 22 | 22 | 22 | 22 | 22 |
| THR33 | EGAD00001003279 | ICGC | EGA | medulloblastoma | 49 | 49 | 49 | 49 | 47 |
| THR35 | EGAD00001004116 | ICR | EGA | high grade glioma | 0 | 1 | 0 | 0 | 0 |
| THR37 | SRP092501 | n/a | SRA | posterior fossa ependymoma | 0 | 0 | 13 | 13 | 13 |
| THR39 | SRP126664 | n/a | SRA | synovial sarcoma | 0 | 0 | 19 | 19 | 19 |
|  |  |  |  | **Repository Sample Sub-total** | **11,195** | **11,248** | **11,300** | **11,963** | **12,352** |
| TH02 | Pacific Pediatric Neuro-Oncology Consortium | Treehouse | Partner | brain cancers | 29 | 29 | 29 | 29 | 29 |
| TH03 | Stanford, Sweet-Cordero Lab | Treehouse | Partner | multiple cancers | 74 | 76 | 76 | 76 | 75 |
| TH04 | Stanford, Sweet-Cordero Lab | Treehouse | Partner | multiple cancers | 1 | 1 | 1 | 1 | 1 |
| TH06 | British Columbia Personalized Onco-Genomics | Treehouse | Partner | multiple cancers | 31 | 39 | 39 | 39 | 39 |
| TH16 | Alberta Children's Hospital, Narendran Lab | Treehouse | Partner | multiple cancers | 0 | 0 | 0 | 2 | 3 |
| TH26 | Stanford - Other | Treehouse | Partner | multiple cancers | 6 | 6 | 6 | 6 | 6 |
| TH27 | UC San Francisco, Sweet-Cordero Lab | Treehouse | Partner | multiple cancers | 29 | 51 | 72 | 79 | 87 |
| TH34 | Stanford Clinical Registry | Treehouse | Partner | multiple cancers | 3 | 6 | 23 | 31 | 35 |
| TH38 | Nationwide Children's Hospital | Treehouse | Partner | multiple cancers | 0 | 0 | 0 | 10 | 10 |
| TH40 | The Hospital for Sick Children | Treehouse | Partner | multiple cancers | 0 | 0 | 0 | 0 | 110 |
|  |  |  |  | **Partner Sample Sub-total** | **173** | **208** | **246** | **273** | **395** |
|  |  |  |  | **Compendium Sample Total** | **11,368** | **11456** | **11,546** | **12,236** | **12,747** |
|  |  |  |  | Number of samples from patients known to be < 30 years old at diagnosis | 1,883 | 1,934 | 1,922 | 2,702 | 2,814 |

#### **Table A6. Size of filtered gene lists for pan-cancer analysis**

| **Treehouse PolyA Compendia Version** | **Number of Expression Filtered Genes (N)** | **Number of Variance Filtered Genes (n)** | **Number of Total Filtered Out Genes (N+n)** | **Number of Genes After Filtering (58581 - (N+n))** |
| --- | --- | --- | --- | --- |
| v7 | 24716 | 6773 | 31489 | 27092 |
| v8 | 24676 | 6781 | 31457 | 27124 |
| v9 | 24726 | 6771 | 31497 | 27084 |
| v10 | 27222 | 6806 | 24553 | 31359 |
| v11 | 24421 | 6832 | 31253 | 27328 |

#### **Table A7. Variants assessed in RNA-Seq data**

| **Chr** | **Start** | **Stop** | **Detectable Mutations** |
| --- | --- | --- | --- |
| chr1 | 114713906 | 114713910 | NRAS c.181C>G (Q61E), c.181C>A (Q61K), c.182A>T (Q61L), c.182A>C (Q61P), c.182A>G (Q61R), c.183A>C (Q61H), c.183A>T (Q61H), c.182_183delAAinsGG (Q61R), c.182_183delAAinsTG (Q61L) |
| chr1 | 114716122 | 114716128 | NRAS c.34G>T (G12C), c.34G>C (G12R), c.34G>A (G12S), c.35G>C (G12A), c.35G>A (G12D), c.35G>T (G12V), c.37G>T (G13C), c.37G>C (G13R), c.38G>C (G13A), c.38G>A (G13D), c.38G>T (G13V) |
| chr1 | 162778599 | 162778601 | DDR2 c.2304T>A (S768R) |
| chr2 | 25234372 | 25234375 | DNMT3A c.2644C>T (R882C), c.2644C>A (R882S), c.2644C>G (R882G), c.2645G>A (R882H), c.2645G>C (R882P), c.2645G>T (R882L) |
| chr2 | 29209788 | 29209790 | ALK c.3833A>C (Y1278S) |
| chr2 | 29209797 | 29209799 | ALK c.3824G>A (R1275Q) |
| chr2 | 29213991 | 29213995 | ALK c.3733T>G (F1245V), c.3734T>G (F1245C), c.3735C>G (F1245L) |
| chr2 | 29214053 | 29214055 | ALK c.3673G>A (D1225N) |
| chr2 | 29220828 | 29220832 | ALK c.3520T>A (F1174I), c.3520T>G (F1174V), c.3521T>G (F1174C), c.3522C>A (F1174L) |
| chr2 | 29222346 | 29222348 | ALK c.3512T>A (I1171N) |
| chr2 | 29222406 | 29222408 | ALK c.3452C>T (T1151M) |
| chr2 | 29223429 | 29223431 | ALK c.3271G>A (D1091N) |
| chr2 | 208248387 | 208248390 | IDH1 c.394C>T (R132C), c.394C>G (R132G), c.394C>A (R132S), c.395G>C (R132P), c.395G>A (R132H), c.395G>T (R132L) |
| chr3 | 41224621 | 41224623 | CTNNB1 c.110C>T (S37F), c.110C>A (S37Y) |
| chr3 | 41224644 | 41224647 | CTNNB1 c.134C>T (S45F), c.134C>A (S45Y), c.133T>C (S45P) |
| chr3 | 179218293 | 179218295 | PIK3CA c.1624G>A (E542K) |
| chr3 | 179218302 | 179218308 | PIK3CA c.1637A>T (Q546L), c.1637A>C (Q546P), c.1637A>G (Q546R), c.1636C>G (Q546E), c.1636C>A (Q546K), c.1634A>G (E545G), c.1634A>T (E545V), c.1633G>C (E545Q), c.1633G>A (E545K) |
| chr3 | 179218314 | 179218316 | PIK3CA c.1645G>A (D549N) |
| chr3 | 179234296 | 179234298 | PIK3CA c.3140A>T (H1047L), c.3140A>G (H1047R) |
| chr4 | 54657926 | 54657928 | KIT c.1730_1738del (P577_D579del), c.1679_1681del (V560del) |
| chr4 | 54725977 | 54725979 | KIT c.1468G>A (E490K) |
| chr4 | 54727424 | 54727426 | KIT c.1657T>A (Y553N) |
| chr4 | 54727436 | 54727438 | KIT c.1669T>A (W557R), c.1669T>C (W557R) |
| chr4 | 54727443 | 54727445 | KIT c.1676T>A (V559D), c.1676T>C (V559A) |
| chr4 | 54727494 | 54727496 | KIT c.1727T>C (L576P) |
| chr4 | 54728054 | 54728056 | KIT c.1924A>G (K642E) |
| chr4 | 54729432 | 54729434 | KIT c.2089C>T (H697Y) |
| chr4 | 54733153 | 54733156 | KIT c.2447A>T (D816V), c.2446G>T (D816Y), c.2446_2447delGAinsAT (D816I), c.2446G>C (D816H) |
| chr4 | 54733167 | 54733169 | KIT c.2460T>A (D820E) |
| chr7 | 55019030 | 55019032 | EGFR c.2290_2291ins (A763_Y764insFQEA) |
| chr7 | 55174013 | 55174016 | EGFR c.2156G>C (G719A), c.2155G>T (G719C), c.2155G>A (G719S) |
| chr7 | 55181377 | 55181379 | EGFR c.2369C>T (T790M) |
| chr7 | 55191821 | 55191823 | EGFR c.2573T>G (L858R) |
| chr7 | 55191830 | 55191832 | EGFR c.2582T>A (L861Q) |
| chr7 | 129189353 | 129189355 | SMO c.203_204delCCinsTT (A68V) |
| chr7 | 129205259 | 129205261 | SMO c.595C>T (R199W) |
| chr7 | 129206274 | 129206276 | SMO c.1046_1047delCCinsTT (T349I) |
| chr7 | 129209347 | 129209349 | SMO c.1417G>C (D473H) |
| chr7 | 129209382 | 129209384 | SMO c.1452_1453delCCinsTT (R485W) |
| chr7 | 129210437 | 129210439 | SMO c.1542_1543delCCinsTT (L515F) |
| chr7 | 129210499 | 129210501 | SMO c.1604G>T (W535L) |
| chr7 | 129210996 | 129210998 | SMO c.1685G>A (R562Q) |
| chr7 | 129212041 | 129212043 | SMO c.1955C>T (A652V) |
| chr7 | 129212349 | 129212351 | SMO c.2263_2264delCCinsTT (P755F) |
| chr7 | 140753333 | 140753338 | BRAF c.1798G>A (V600M), c.1799T>G (V600G), c.1798_1799delGTinsAA (V600K), c.1798_1799delGTinsAG (V600R), c.1799T>A (V600E), c.1799_1800delTGinsAA (V600E), c.1799_1800delTGinsAT (V600D), c.1801A>G (K601E) |
| chr7 | 140753344 | 140753347 | BRAF c.1789C>G (L597V), c.1790T>A (L597Q), c.1789_1790delCTinsTC (L597S), c.1790T>G (L597R) |
| chr7 | 140753348 | 140753350 | BRAF c.1786G>C (G596R) |
| chr7 | 140753352 | 140753356 | BRAF c.1780G>A (D594N), c.1779_1780delTGinsGA (D594N), c.1780G>C (D594H), c.1781A>G (D594G), c.1781A>T (D594V), c.1782T>A (D594E), c.1782T>G (D594E) |
| chr7 | 140781592 | 140781594 | BRAF c.1415A>G (Y472C) |
| chr7 | 140781601 | 140781603 | BRAF c.1406G>A (G469E), c.1406G>T (G469V), c.1405_1406delGGinsTT (G469L), c.1406G>C (G469A) |
| chr7 | 140781610 | 140781612 | BRAF c.1397G>T (G466V) |
| chr9 | 5078359 | 5078363 | JAK2 c.2049A>T (R683S), c.2049A>C (R683S), c.2048G>C (R683T), c.2047A>G (R683G) |
| chr9 | 77794571 | 77794573 | GNAQ c.626A>T (Q209L), c.626A>C (Q209P), c.626A>G (Q209R) |
| chr9 | 77797576 | 77797578 | GNAQ c.548G>A (R183Q) |
| chr9 | 132891346 | 132891348 | TSC1 c.1907_1908del (E636fs) |
| chr9 | 136496196 | 136496198 | NOTCH1 c.7541_7542delCT (P2514fs) |
| chr10 | 87933146 | 87933149 | PTEN c.389delG (R130fs*4), c.389G>A (R130Q), c.388C>T (R130*), c.388C>G (R130G) |
| chr10 | 87933235 | 87933237 | PTEN c.477G>T (R159S) |
| chr10 | 87957914 | 87957916 | PTEN c.697C>T (R233*) |
| chr10 | 87957958 | 87957960 | PTEN c.741dupA (P248fs*5) |
| chr10 | 87958017 | 87958019 | PTEN c.800delA (K267fs*9) |
| chr10 | 87961059 | 87961061 | PTEN c.968dupA (N323fs*2), c.968delA (N323fs*21) |
| chr11 | 533873 | 533875 | HRAS c.182A>G (Q61R) |
| chr11 | 534285 | 534290 | HRAS c.34G>C (G12R), c.35G>T (G12V), c.37G>T (G13C), c.37G>C (G13R) |
| chr12 | 25225626 | 25225629 | KRAS c.436G>C (A146P), c.436G>A (A146T), c.437C>T (A146V) |
| chr12 | 25225712 | 25225714 | KRAS c.351A>C (K117N), c.351A>T (K117N) |
| chr12 | 25227340 | 25227344 | KRAS c.181C>A (Q61K), c.182A>C (Q61P), c.182A>T (Q61L), c.182A>G (Q61R), c.183A>C (Q61H), c.183A>T (Q61H) |
| chr12 | 25245346 | 25245352 | KRAS c.34G>T (G12C), c.34G>C (G12R), c.34G>A (G12S), c.35G>C (G12A), c.35G>A (G12D), c.35G>T (G12V), c.37G>T (G13C), c.37G>C (G13R), c.37G>A (G13S), c.38G>T (G13V), c.38G>C (G13A), c.38G>A (G13D) |
| chr13 | 28018499 | 28018506 | FLT3 c.2503G>C (D835H), c.2503G>A (D835N), c.2503G>T (D835Y), c.2504A>C (D835A), c.2504A>T (D835V), c.2505T>G (D835E), c.2505T>A (D835E), c.2506A>C (I836L), c.2506A>G (I836V), c.2506A>T (I836F), c.2506_2507delATinsGA (I836D), c.2506_2507delATinsCA (I836H), c.2508C>G (I836M) |
| chr14 | 104780213 | 104780215 | AKT1 c.49G>A (E17K) |
| chr15 | 66435102 | 66435104 | MEK1 c.157T>C (F53L) |
| chr15 | 66435112 | 66435114 | MEK1 c.167A>C (Q56P) |
| chr15 | 66435116 | 66435118 | MEK1 c.171G>T (K57N) |
| chr15 | 66435144 | 66435146 | MEK1 c.199G>A (D67N) |
| chr15 | 66436785 | 66436787 | MEK1 c.332T>G (I111S) |
| chr15 | 66436815 | 66436817 | MEK1 c.362G>C (C121S) |
| chr15 | 66436823 | 66436826 | MEK1 c.371C>T (P124L), c.370C>T (P124S) |
| chr15 | 66481792 | 66481794 | MEK1 c.607G>A (E203K) |
| chr15 | 66485085 | 66485087 | MEK1 c.790C>T (P264S) |
| chr15 | 66490576 | 66490578 | MEK1 c.1144A>C (N382H) |
| chr15 | 90088604 | 90088608 | IDH2 c.514A>G (R172G), c.514A>T (R172W), c.515G>A (R172K), c.515G>T (R172M), c.516G>C (R172S), c.516G>T (R172S) |
| chr15 | 90088701 | 90088704 | IDH2 c.418C>T (R140W), c.418C>G (R140G), c.419G>A (R140Q), c.419G>T (R140L) |
| chr17 | 39688082 | 39688084 | HER2 c.2339_2340ins (G778_P780dup) |
| chr17 | 39711951 | 39711953 | HER2 c.926G>C (G309A) |
| chr17 | 39723966 | 39723968 | HER2 c.2264_2278del (L755_T759del), c.2264T>C (L755S) |
| chr17 | 39724007 | 39724009 | HER2 c.2305G>C (D769H), c.2305G>T (D769Y) |
| chr17 | 39724746 | 39724748 | HER2 c.2329G>T (V777L) |
| chr17 | 39725078 | 39725080 | HER2 c.2524G>A (V842I) |
| chr17 | 39725362 | 39725364 | HER2 c.2686C>T (R896C) |
| chr18 | 51065455 | 51065457 | SMAD4 c.989A>C (E330A) |
| chr18 | 51065517 | 51065519 | SMAD4 c.1051G>C (D351H), c.1051G>A (D351N) |
| chr18 | 51065531 | 51065533 | SMAD4 c.1065C>A (D355E) |
| chr18 | 51065547 | 51065550 | SMAD4 c.1082G>A (R361H), c.1081C>A (R361S), c.1081C>T (R361C) |
| chr18 | 51078416 | 51078418 | SMAD4 c.1609G>T (D537Y) |
| chr19 | 3115012 | 3115015 | GNA11 c.547C>T (R183C), c.546_547delCCinsTT (R183C) |
| chr19 | 3118943 | 3118945 | GNA11 c.626A>C (Q209P), c.626A>T (Q209L) |

**Table A8. Known cancer fusion genes**

| ABI1 | C11orf95 | CTNNB1 | FGF1 | HOOK3 | LCP1 | MYH11 | PAX5 | PTPRK | SH3GL1 | TFRC |
| --- | --- | --- | --- | --- | --- | --- | --- | --- | --- | --- |
| ABL1 | CAMTA1 | DDIT3 | FGFR1 | HOXA11 | LIFR | MYH9 | PAX7 | PVT1 | SLC1A2 | TGFBR3 |
| ABL2 | CARS | DDX10 | FGFR1OP | HOXA13 | LMO1 | MYST3 | PAX8 | RABEP1 | SNX29 | THADA |
| ACSL6 | CBFA2T3 | DDX6 | FGFR2 | HOXA3 | LMO2 | NACA | PBX1 | RAF1 | SPTAN1 | TLX1 |
| AF10 | CBFB | DEK | FGFR3 | HOXA9 | LPP | NBEAP1 | PCM1 | RALGDS | SRSF3 | TLX3 |
| AF6 | CBL | DNAJB1 | FGFR4 | HOXC11 | LYL1 | NCOA2 | PCSK7 | RANBP17 | SS18 | TMPRSS2 |
| AFF1 | CCND1 | DUSP22 | FLI1 | HOXC13 | MAF | NDRG1 | PDE4DIP | RANBP2 | SSBP2 | TNFRSF11A |
| AFF4 | CCND2 | E2A | FN1 | HOXD11 | MAFB | NF1 | PDGFB | RAP1GDS1 | SSX1 | TNIP1 |
| ALK | CCND3 | EBF1 | FNBP1 | HOXD13 | MALT1 | NF2 | PDGFRA | RARA | SSX2 | TOP1 |
| ARHGAP26 | CD274 | EGFR | FOSB1 | HSP90AA1 | MAML2 | NFKB2 | PDGFRB | RBM15 | SSX4 | TP53 |
| ARHGEF12 | CD74 | EIF4A2 | FOXO1 | HSP90AB1 | MDS2 | NIN | PDL1 | RCSD1 | STAT6 | TP63 |
| ARID1A | CDK6 | ELF4 | FOXO3 | IGH | MECOM | NOTCH1 | PER1 | RELA | STL | TPM3 |
| ARNT | CDX2 | ELL | FOXO4 | IGK | MET | NOTCH2 | PHF1 | RET | STRN3 | TPM4 |
| ASXL1 | CENPC | ELN | FOXP1 | IGL | MGEA5 | NPM1 | PICALM | RFX3 | SYK | TRIM24 |
| ATF1 | CEP110 | EML4 | FOXR1 | IKZF1 | MKL1 | NR4A3 | PIM1 | RHOH | TAF15 | TRIP11 |
| ATG5 | CHIC2 | ENL | FOXR2 | IL21R | MKL2 | NSD1 | PLAG1 | RNF213 | TAL1 | TTL |
| ATIC | CHN1 | EP300 | FSTL3 | IL3 | MLF1 | NTRK1 | PML | ROS1 | TAL2 | TYK2 |
| BCL10 | CIC | EPOR | FUS | IRF4 | MLL | NTRK2 | POU2AF1 | ROS1 | TBL1XR1 | USP6 |
| BCL11A | CIITA | EPS15 | GAB2 | ITK | MLLT1 | NTRK3 | PPARG | RPL22 | TCF3 | WHSC1L1 |
| BCL11B | CLP1 | ERBB2 | GAS7 | JAK1 | MLLT10 | NUMA1 | PPP1CB | RPN1 | TCF3 | WT1 |
| BCL2 | CLTC | ERCC2 | GLI1 | JAK2 | MLLT3 | NUP153 | PRDM1 | RSPO2 | TCF7L2 | WTR1 |
| BCL3 | CLTCL1 | ERG | GMPS | JAK3 | MLLT4 | NUP160 | PRDM16 | RSPO3 | TCL1 | YAP1 |
| BCL6 | CNTRL | ETS1 | GPHN | JAZF1 | MLLT6 | NUP214 | PRKACA | RUNDC2A | TCL1A | YPEL5 |
| BCL7A | COL1A1 | ETV1 | GRM1 | KAT6A | MN1 | NUP98 | PRKAR1A | RUNX1 | TEC | YWHAE |
| BCL9 | CREB1 | ETV4 | HERPUD1 | KDSR | MNX1 | NUTM1 | PRKCA | RUNX2 | TERF2 | ZBTB16 |
| BCOR | CREB3L1 | ETV5 | HEY1 | KIF5B | MSI2 | NUTM2A | PRKCB | SEC31A | TERT | ZMIZ1 |
| BCR | CREB3L2 | ETV6 | HIP1 | KMT2A | MSN | OMD | PRKCD | SEPT5 | TET1 | ZMYM2 |
| BIRC3 | CREBBP | EWSR1 | HIST1H4I | KMT2A | MUC1 | P2RY8 | PRRX1 | SEPT6 | TFE3 | ZNF274 |
| BRAF | CRLF2 | FCGR2B | HLF | KMT2C | MYB | PAFAH1B2 | PSIP1 | SEPT9 | TFEB | ZNF384 |
| BRD4 | CSF1 | FCRL4 | HMGA1 | KRAS | MYBL1 | PAG1 | PTCH1 | SET | TFG | ZNF521 |
| BTG1 | CSF1R | FEV | HMGA2 | LASP1 | MYC | PAX3 | PTK7 | SFPQ | TFPT | ZNF703 |

#### **Table A9. Key registry timepoints per sample**

| **Sample ID** | **Collection to Shipping (Days)** | **Shipping to Arrival at Covance (Days)** | **Extract Prep Sequence (Days)** | **Automated Treehouse Analysis (Days)** | **Mock to TB (Days)** | **Overall Turnaround (Days)** |
| --- | --- | --- | --- | --- | --- | --- |
| TH34_1162_S01 | 13 | 1 | 12 | 3 | 13 | 34 |
| TH34_1163_S01 | 5 | 1 | 24 | 4 | 1 | 38 |
| TH34_1149_S02 | 7 | 1 | 8 | 2 | 2 | 23 |
| TH34_1179_S01 | 18 | 1 | 7 | 1 | 11 | 29 |
| TH34_1238_S01 | 1 | 1 | 5 | 3 | 3 | 14 |
| TH34_1239_S01 | 3 | 1 | 4 | 1 | 3 | 18 |
| TH34_1240_S01 | 1 | 1 | 4 | 1 | 1 | 8 |
| TH34_1349_S01 | 2 | 1 | 4 | 4 | 2 | 20 |
| TH34_1349_S02 | 2 | 1 | 4 | 4 | 2 | 20 |
| TH34_1350_S01 | 1 | 1 | 5 | 13 | 3 | 25 |
| TH34_1351_S01 | 2 | 1 | 5 | 5 | 3 | 18 |
| TH34_1352_S01 | 1 | 1 | 3 | 2 | 3 | 11 |
| TH34_1379_S01 | 2 | 1 | 4 | 2 | 3 | 11 |
| TH34_1380_S01 | 1 | 1 | 7 | 2 | 2 | 16 |
| TH34_1381_S01 | 226 | 1 | 7 | 5 | 3 | 241 |
| TH34_1150_S02 | 1 | 1 | 7 | 5 | 3 | 16 |
| TH34_1399_S01 | 3 | 1 | 7 | 2 | 2 | 15 |
| TH34_1400_S01 | 2 | 1 | 7 | 5 | 12 | 18 |
| TH34_1412_S01 | 2 | 1 | 6 | 1 | 3 | 15 |
| TH34_1414_S01 | 4 | 1 | 8 | 2 | 3 | 19 |
| TH34_1415_S01 | 2 | 1 | 8 | 2 | 4 | 16 |
| TH34_1444_S01 | 2 | 1 | 5 | 2 | 3 | 12 |
| TH34_1445_S02 | 17 | 1 | 8 | 4 | 3 | 32 |
| TH34_1446_S01 | 2 | 1 | 4 | 3 | 3 | 11 |
| TH34_1447_S01 | 0 | 1 | 4 | 4 | 3 | 21 |
| TH34_1447_S02 | 1 | 1 | 3 | 4 | 3 | 21 |
| TH34_1452_S01 | 2 | 1 | 6 | 2 | 3 | 20 |
| TH34_1455_S01 | 1 | 1 | 7 | 1 | 3 | 25 |
| TH34_1456_S02 | 2 | 1 | 14 | 4 | 3 | 25 |
| TH34_2292_S01 | 3 | 1 | 7 | 2 | 3 | 14 |
| TH34_2293_S01 | 1 | 1 | 4 | 1 | 2 | 12 |
| TH34_2351_S01 | 9 | 1 | 4 | 1 | 1 | 18 |
| TH34_2410_S01 | 13 | 1 | 7 | 5 | 7 | 29 |
| TH34_2411_S01 | 4 | 1 | 7 | 5 | 7 | 20 |
| TH34_2666_S01 | 10 | 1 | 16 | 2 | 5 | 33 |

#### **Table A10. Outliers detected for each dataset**

| **Sample ID** | **Outlier gene** | **Comparison cohort** | **Pathway support** |
| --- | --- | --- | --- |
| TH34_1149_S02 | IGF1 | Pediatric | TRUE |
| TH34_1149_S02 | IGF1 | Stanford | TRUE |
| TH34_1149_S02 | IGF1 | TCGA | TRUE |
| TH34_1149_S02 | IGF1 | Treehouse pan-cancer | TRUE |
| TH34_1149_S02 | IGF1 | Treehouse pan-disease | FALSE |
| TH34_1149_S02 | NTRK3 | TCGA | FALSE |
| TH34_1149_S02 | NTRK3 | Treehouse pan-cancer | FALSE |
| TH34_1149_S02 | NTRK3 | Treehouse pan-disease | FALSE |
| TH34_1150_S02 | BCL6 | Treehouse pan-disease | FALSE |
| TH34_1150_S02 | CCND1 | Stanford | TRUE |
| TH34_1150_S02 | CCND1 | TCGA | TRUE |
| TH34_1150_S02 | CDK9 | Treehouse pan-disease | FALSE |
| TH34_1150_S02 | JAK1 | Pediatric | FALSE |
| TH34_1150_S02 | JAK1 | Stanford | FALSE |
| TH34_1150_S02 | JAK1 | TCGA | FALSE |
| TH34_1150_S02 | JAK1 | Treehouse pan-cancer | FALSE |
| TH34_1150_S02 | PDCD1 | Pediatric | FALSE |
| TH34_1150_S02 | PDCD1 | Stanford | FALSE |
| TH34_1150_S02 | PDCD1 | TCGA | FALSE |
| TH34_1150_S02 | PDCD1 | Treehouse pan-cancer | FALSE |
| TH34_1162_S01 | CSF1R | Treehouse pan-disease | TRUE |
| TH34_1162_S01 | GATA2 | TCGA | FALSE |
| TH34_1162_S01 | HMOX1 | Pediatric | FALSE |
| TH34_1162_S01 | HMOX1 | TCGA | FALSE |
| TH34_1162_S01 | HMOX1 | Treehouse pan-cancer | FALSE |
| TH34_1162_S01 | HMOX1 | Treehouse pan-disease | TRUE |
| TH34_1162_S01 | JAK1 | Stanford | FALSE |
| TH34_1162_S01 | JAK1 | TCGA | FALSE |
| TH34_1162_S01 | JAK1 | Treehouse pan-cancer | FALSE |
| TH34_1163_S01 | IGF2 | TCGA | FALSE |
| TH34_1163_S01 | IGF2 | Treehouse pan-cancer | FALSE |
| TH34_1179_S01 | FGFR1 | TCGA | TRUE |
| TH34_1179_S01 | FGFR1 | Treehouse pan-cancer | TRUE |
| TH34_1179_S01 | PTCH1 | TCGA | TRUE |
| TH34_1179_S01 | PTCH1 | Treehouse pan-cancer | TRUE |
| TH34_1238_S01 | BTK | Stanford | TRUE |
| TH34_1238_S01 | BTK | TCGA | TRUE |
| TH34_1238_S01 | BTK | Treehouse pan-cancer | TRUE |
| TH34_1238_S01 | CCND3 | TCGA | TRUE |
| TH34_1238_S01 | CDK9 | TCGA | TRUE |
| TH34_1238_S01 | MS4A1 | Pediatric | FALSE |
| TH34_1238_S01 | MS4A1 | Stanford | TRUE |
| TH34_1238_S01 | MS4A1 | TCGA | TRUE |
| TH34_1238_S01 | MS4A1 | Treehouse pan-cancer | TRUE |
| TH34_1238_S01 | MTOR | Stanford | TRUE |
| TH34_1238_S01 | MTOR | TCGA | TRUE |
| TH34_1238_S01 | PIK3CD | Stanford | TRUE |
| TH34_1238_S01 | PIK3CD | TCGA | TRUE |
| TH34_1238_S01 | PIK3CD | Treehouse pan-cancer | TRUE |
| TH34_1238_S01 | RPTOR | Stanford | TRUE |
| TH34_1238_S01 | RPTOR | TCGA | TRUE |
| TH34_1239_S01 | BTK | TCGA | TRUE |
| TH34_1239_S01 | BTK | Treehouse pan-cancer | TRUE |
| TH34_1239_S01 | CCND3 | TCGA | TRUE |
| TH34_1239_S01 | PIK3CD | Stanford | TRUE |
| TH34_1239_S01 | PIK3CD | TCGA | TRUE |
| TH34_1239_S01 | PIK3CD | Treehouse pan-cancer | TRUE |
| TH34_1239_S01 | PIK3R5 | TCGA | TRUE |
| TH34_1239_S01 | PIK3R5 | Treehouse pan-cancer | TRUE |
| TH34_1349_S01 | ETV1 | TCGA | FALSE |
| TH34_1349_S01 | ETV1 | Treehouse pan-cancer | TRUE |
| TH34_1349_S01 | IGF2 | TCGA | TRUE |
| TH34_1349_S01 | IGF2 | Treehouse pan-cancer | TRUE |
| TH34_1349_S01 | KIT | Pediatric | FALSE |
| TH34_1349_S01 | KIT | Stanford | FALSE |
| TH34_1349_S01 | KIT | TCGA | FALSE |
| TH34_1349_S01 | KIT | Treehouse pan-cancer | FALSE |
| TH34_1349_S01 | PTCH1 | TCGA | TRUE |
| TH34_1349_S02 | ETV1 | TCGA | TRUE |
| TH34_1349_S02 | ETV1 | Treehouse pan-cancer | TRUE |
| TH34_1349_S02 | IGF2 | TCGA | TRUE |
| TH34_1349_S02 | KIT | Pediatric | FALSE |
| TH34_1349_S02 | KIT | Stanford | FALSE |
| TH34_1349_S02 | KIT | TCGA | FALSE |
| TH34_1349_S02 | KIT | Treehouse pan-cancer | FALSE |
| TH34_1349_S02 | PTCH1 | TCGA | TRUE |
| TH34_1349_S02 | PTCH1 | Treehouse pan-cancer | TRUE |
| TH34_1350_S01 | BTK | TCGA | TRUE |
| TH34_1350_S01 | BTK | Treehouse pan-cancer | TRUE |
| TH34_1350_S01 | PIK3CD | TCGA | TRUE |
| TH34_1350_S01 | PIK3R5 | TCGA | TRUE |
| TH34_1350_S01 | PIK3R5 | Treehouse pan-cancer | TRUE |
| TH34_1351_S01 | MAP2K4 | Stanford | FALSE |
| TH34_1351_S01 | MAP2K4 | TCGA | FALSE |
| TH34_1351_S01 | MAP2K4 | Treehouse pan-cancer | FALSE |
| TH34_1351_S01 | MAP2K4 | Treehouse pan-disease | TRUE |
| TH34_1352_S01 | CCND2 | Treehouse pan-disease | FALSE |
| TH34_1352_S01 | FGFR1 | TCGA | TRUE |
| TH34_1352_S01 | FGFR1 | Treehouse pan-cancer | TRUE |
| TH34_1352_S01 | FGFR2 | TCGA | TRUE |
| TH34_1352_S01 | IGF2 | TCGA | TRUE |
| TH34_1352_S01 | IGF2 | Treehouse pan-cancer | TRUE |
| TH34_1352_S01 | PDGFRA | TCGA | TRUE |
| TH34_1379_S01 | CCND2 | Treehouse pan-disease | TRUE |
| TH34_1379_S01 | FGFR4 | Pediatric | FALSE |
| TH34_1379_S01 | HMOX1 | TCGA | TRUE |
| TH34_1379_S01 | HMOX1 | Treehouse pan-cancer | TRUE |
| TH34_1379_S01 | HMOX1 | Treehouse pan-disease | TRUE |
| TH34_1379_S01 | IGF2 | TCGA | TRUE |
| TH34_1379_S01 | IGF2 | Treehouse pan-disease | TRUE |
| TH34_1380_S01 | FGFR4 | Pediatric | FALSE |
| TH34_1380_S01 | IGF2 | TCGA | FALSE |
| TH34_1380_S01 | IGF2 | Treehouse pan-cancer | TRUE |
| TH34_1380_S01 | MAP2K2 | Stanford | TRUE |
| TH34_1380_S01 | NOTCH3 | Stanford | TRUE |
| TH34_1380_S01 | NOTCH3 | TCGA | TRUE |
| TH34_1380_S01 | NOTCH3 | Treehouse pan-cancer | TRUE |
| TH34_1380_S01 | NOTCH3 | Treehouse pan-disease | TRUE |
| TH34_1381_S01 | HSP90B1 | Stanford | TRUE |
| TH34_1381_S01 | HSP90B1 | TCGA | TRUE |
| TH34_1381_S01 | HSP90B1 | Treehouse pan-cancer | TRUE |
| TH34_1381_S01 | HSP90B1 | Treehouse pan-disease | TRUE |
| TH34_1381_S01 | IGF2 | TCGA | TRUE |
| TH34_1381_S01 | NTRK2 | Stanford | TRUE |
| TH34_1381_S01 | PARP1 | Treehouse pan-disease | FALSE |
| TH34_1381_S01 | PIK3R1 | Stanford | TRUE |
| TH34_1381_S01 | TSC2 | TCGA | TRUE |
| TH34_1381_S01 | VEGFA | Stanford | TRUE |
| TH34_1381_S01 | VEGFA | TCGA | TRUE |
| TH34_1381_S01 | VEGFA | Treehouse pan-cancer | TRUE |
| TH34_1399_S01 | IGF1 | Pediatric | TRUE |
| TH34_1399_S01 | IGF1 | TCGA | TRUE |
| TH34_1399_S01 | IGF1 | Treehouse pan-cancer | TRUE |
| TH34_1399_S01 | IGF2 | TCGA | TRUE |
| TH34_1399_S01 | IGF2 | Treehouse pan-cancer | TRUE |
| TH34_1400_S01 | IGF1 | Pediatric | TRUE |
| TH34_1400_S01 | NTRK2 | Stanford | TRUE |
| TH34_1400_S01 | STAT5A | Treehouse pan-disease | TRUE |
| TH34_1412_S01 | FGFR4 | Pediatric | TRUE |
| TH34_1412_S01 | IGF2 | TCGA | TRUE |
| TH34_1412_S01 | IGF2 | Treehouse pan-cancer | TRUE |
| TH34_1412_S01 | KDR | Stanford | TRUE |
| TH34_1412_S01 | KDR | TCGA | TRUE |
| TH34_1412_S01 | KDR | Treehouse pan-cancer | TRUE |
| TH34_1412_S01 | NTRK3 | TCGA | TRUE |
| TH34_1412_S01 | NTRK3 | Treehouse pan-cancer | TRUE |
| TH34_1414_S01 | ALK | Pediatric | FALSE |
| TH34_1414_S01 | ALK | Stanford | FALSE |
| TH34_1414_S01 | ALK | TCGA | FALSE |
| TH34_1414_S01 | ALK | Treehouse pan-cancer | FALSE |
| TH34_1414_S01 | CDK4 | Stanford | TRUE |
| TH34_1414_S01 | CDK4 | TCGA | TRUE |
| TH34_1414_S01 | CDK4 | Treehouse pan-cancer | TRUE |
| TH34_1414_S01 | FGFR4 | Pediatric | FALSE |
| TH34_1414_S01 | FGFR4 | Treehouse pan-cancer | FALSE |
| TH34_1414_S01 | IGF2 | TCGA | TRUE |
| TH34_1414_S01 | IGF2 | Treehouse pan-cancer | FALSE |
| TH34_1414_S01 | STAT2 | Stanford | FALSE |
| TH34_1414_S01 | STAT2 | TCGA | FALSE |
| TH34_1414_S01 | STAT2 | Treehouse pan-cancer | FALSE |
| TH34_1414_S01 | STAT2 | Treehouse pan-disease | FALSE |
| TH34_1415_S01 | FLT4 | Stanford | FALSE |
| TH34_1415_S01 | FLT4 | TCGA | FALSE |
| TH34_1415_S01 | FLT4 | Treehouse pan-cancer | FALSE |
| TH34_1415_S01 | FLT4 | Treehouse pan-disease | FALSE |
| TH34_1415_S01 | HMOX1 | Pediatric | TRUE |
| TH34_1415_S01 | HMOX1 | TCGA | TRUE |
| TH34_1415_S01 | HMOX1 | Treehouse pan-cancer | TRUE |
| TH34_1415_S01 | HMOX1 | Treehouse pan-disease | FALSE |
| TH34_1415_S01 | IL6 | Pediatric | TRUE |
| TH34_1415_S01 | IL6 | Stanford | TRUE |
| TH34_1415_S01 | IL6 | TCGA | TRUE |
| TH34_1415_S01 | IL6 | Treehouse pan-cancer | TRUE |
| TH34_1444_S01 | AKT1 | Stanford | TRUE |
| TH34_1444_S01 | AKT1 | Treehouse pan-disease | TRUE |
| TH34_1444_S01 | AKT2 | Pediatric | TRUE |
| TH34_1444_S01 | AKT2 | Stanford | TRUE |
| TH34_1444_S01 | AKT2 | TCGA | TRUE |
| TH34_1444_S01 | AKT2 | Treehouse pan-cancer | TRUE |
| TH34_1444_S01 | AKT2 | Treehouse pan-disease | TRUE |
| TH34_1444_S01 | DEPTOR | Stanford | TRUE |
| TH34_1444_S01 | KIT | Pediatric | TRUE |
| TH34_1444_S01 | KIT | Stanford | TRUE |
| TH34_1444_S01 | KIT | TCGA | TRUE |
| TH34_1444_S01 | KIT | Treehouse pan-cancer | TRUE |
| TH34_1444_S01 | KIT | Treehouse pan-disease | TRUE |
| TH34_1444_S01 | NTRK2 | Stanford | TRUE |
| TH34_1444_S01 | PIK3R2 | Treehouse pan-disease | TRUE |
| TH34_1444_S01 | RAF1 | Treehouse pan-disease | TRUE |
| TH34_1444_S01 | TSC2 | Stanford | TRUE |
| TH34_1444_S01 | TSC2 | TCGA | TRUE |
| TH34_1444_S01 | TSC2 | Treehouse pan-cancer | TRUE |
| TH34_1444_S01 | TSC2 | Treehouse pan-disease | TRUE |
| TH34_1444_S01 | VEGFC | Pediatric | TRUE |
| TH34_1444_S01 | VEGFC | Stanford | TRUE |
| TH34_1444_S01 | VEGFC | TCGA | TRUE |
| TH34_1444_S01 | VEGFC | Treehouse pan-cancer | TRUE |
| TH34_1444_S01 | VEGFC | Treehouse pan-disease | TRUE |
| TH34_1445_S02 | ETV1 | TCGA | FALSE |
| TH34_1445_S02 | HMOX1 | Treehouse pan-disease | TRUE |
| TH34_1445_S02 | IGF2 | TCGA | TRUE |
| TH34_1445_S02 | IGF2 | Treehouse pan-disease | TRUE |
| TH34_1445_S02 | NTRK2 | Stanford | TRUE |
| TH34_1445_S02 | NTRK3 | Stanford | TRUE |
| TH34_1445_S02 | NTRK3 | TCGA | FALSE |
| TH34_1445_S02 | NTRK3 | Treehouse pan-cancer | FALSE |
| TH34_1445_S02 | VEGFA | TCGA | TRUE |
| TH34_1445_S02 | VEGFA | Treehouse pan-disease | TRUE |
| TH34_1445_S02 | WEE1 | Stanford | FALSE |
| TH34_1445_S02 | WEE1 | TCGA | TRUE |
| TH34_1445_S02 | WEE1 | Treehouse pan-cancer | TRUE |
| TH34_1445_S02 | WEE1 | Treehouse pan-disease | TRUE |
| TH34_1446_S01 | NTRK2 | Stanford | TRUE |
| TH34_1446_S01 | NTRK3 | TCGA | FALSE |
| TH34_1446_S01 | NTRK3 | Treehouse pan-cancer | FALSE |
| TH34_1447_S01 | CCNE1 | Treehouse pan-disease | TRUE |
| TH34_1447_S01 | HMOX1 | TCGA | TRUE |
| TH34_1447_S01 | IGF2 | TCGA | TRUE |
| TH34_1447_S01 | PARP2 | TCGA | TRUE |
| TH34_1447_S01 | PARP2 | Treehouse pan-cancer | TRUE |
| TH34_1447_S01 | PARP2 | Treehouse pan-disease | TRUE |
| TH34_1447_S02 | HMOX1 | TCGA | TRUE |
| TH34_1447_S02 | HMOX1 | Treehouse pan-cancer | FALSE |
| TH34_1447_S02 | IGF2 | TCGA | FALSE |
| TH34_1447_S02 | IGF2 | Treehouse pan-cancer | FALSE |
| TH34_1447_S02 | PARP2 | TCGA | TRUE |
| TH34_1452_S01 | AKT1 | Stanford | TRUE |
| TH34_1452_S01 | FGFR3 | Pediatric | FALSE |
| TH34_1452_S01 | FGFR3 | Stanford | TRUE |
| TH34_1452_S01 | FGFR3 | TCGA | TRUE |
| TH34_1452_S01 | HDAC4 | Pediatric | FALSE |
| TH34_1452_S01 | HDAC4 | Stanford | TRUE |
| TH34_1452_S01 | HDAC4 | TCGA | TRUE |
| TH34_1452_S01 | HDAC4 | Treehouse pan-cancer | FALSE |
| TH34_1452_S01 | HDAC4 | Treehouse pan-disease | TRUE |
| TH34_1452_S01 | IGF2 | TCGA | TRUE |
| TH34_1452_S01 | IGF2 | Treehouse pan-cancer | TRUE |
| TH34_1452_S01 | NTRK2 | Stanford | TRUE |
| TH34_1452_S01 | PIK3R2 | Stanford | TRUE |
| TH34_1452_S01 | PIK3R2 | TCGA | TRUE |
| TH34_1452_S01 | PIK3R2 | Treehouse pan-cancer | TRUE |
| TH34_1452_S01 | TSC2 | Stanford | TRUE |
| TH34_1452_S01 | TSC2 | TCGA | TRUE |
| TH34_1452_S01 | TSC2 | Treehouse pan-cancer | TRUE |
| TH34_1455_S01 | CDK9 | Stanford | FALSE |
| TH34_1455_S01 | CDK9 | TCGA | TRUE |
| TH34_1455_S01 | CDK9 | Treehouse pan-cancer | FALSE |
| TH34_1455_S01 | FGFR3 | TCGA | FALSE |
| TH34_1455_S01 | IGF2 | TCGA | FALSE |
| TH34_1455_S01 | IGF2 | Treehouse pan-cancer | FALSE |
| TH34_1455_S01 | NTRK2 | Stanford | FALSE |
| TH34_1455_S01 | NTRK2 | Treehouse pan-cancer | FALSE |
| TH34_1456_S02 | GATA2 | TCGA | TRUE |
| TH34_1456_S02 | GATA2 | Treehouse pan-cancer | FALSE |
| TH34_1456_S02 | GATA2 | Treehouse pan-disease | TRUE |
| TH34_1456_S02 | HDAC7 | Treehouse pan-disease | TRUE |
| TH34_1456_S02 | KDR | TCGA | TRUE |
| TH34_2292_S01 | BCL6 | TCGA | TRUE |
| TH34_2292_S01 | ETV1 | TCGA | FALSE |
| TH34_2292_S01 | FLT4 | Pediatric | TRUE |
| TH34_2292_S01 | FLT4 | Stanford | TRUE |
| TH34_2292_S01 | FLT4 | TCGA | TRUE |
| TH34_2292_S01 | FLT4 | Treehouse pan-cancer | TRUE |
| TH34_2292_S01 | HMOX1 | TCGA | TRUE |
| TH34_2292_S01 | HMOX1 | Treehouse pan-cancer | TRUE |
| TH34_2292_S01 | VEGFA | Stanford | TRUE |
| TH34_2292_S01 | VEGFA | TCGA | TRUE |
| TH34_2292_S01 | VEGFA | Treehouse pan-cancer | TRUE |
| TH34_2293_S01 | IGF2 | TCGA | TRUE |
| TH34_2293_S01 | IGF2 | Treehouse pan-cancer | TRUE |
| TH34_2293_S01 | STAT1 | Stanford | FALSE |
| TH34_2351_S01 | FGFR4 | Pediatric | FALSE |
| TH34_2351_S01 | HMOX1 | Treehouse pan-disease | TRUE |
| TH34_2351_S01 | IGF2 | TCGA | TRUE |
| TH34_2351_S01 | IGF2 | Treehouse pan-cancer | TRUE |
| TH34_2410_S01 | IGF2 | TCGA | TRUE |
| TH34_2411_S01 | CDK4 | Pediatric | TRUE |
| TH34_2411_S01 | CDK4 | Stanford | TRUE |
| TH34_2411_S01 | CDK4 | TCGA | TRUE |
| TH34_2411_S01 | CDK4 | Treehouse pan-cancer | TRUE |
| TH34_2411_S01 | CDK4 | Treehouse pan-disease | TRUE |
| TH34_2411_S01 | FGFR1 | TCGA | TRUE |
| TH34_2411_S01 | MAP2K2 | Stanford | TRUE |
| TH34_2411_S01 | MAP2K2 | TCGA | TRUE |
| TH34_2411_S01 | MAP2K2 | Treehouse pan-cancer | TRUE |
| TH34_2411_S01 | MAP2K2 | Treehouse pan-disease | TRUE |
| TH34_2411_S01 | MDM2 | Pediatric | TRUE |
| TH34_2411_S01 | MDM2 | Stanford | TRUE |
| TH34_2411_S01 | MDM2 | TCGA | TRUE |
| TH34_2411_S01 | MDM2 | Treehouse pan-cancer | TRUE |
| TH34_2411_S01 | MDM2 | Treehouse pan-disease | TRUE |
| TH34_2666_S01 | FLT4 | TCGA | FALSE |
| TH34_2666_S01 | FLT4 | Treehouse pan-cancer | FALSE |
| TH34_2666_S01 | MDM2 | Pediatric | FALSE |
| TH34_2666_S01 | MDM2 | Stanford | FALSE |
| TH34_2666_S01 | MDM2 | TCGA | FALSE |
| TH34_2666_S01 | MDM2 | Treehouse pan-cancer | FALSE |

#### **Table A11. Cohort-specific features of expression distributions for TCGA-only outlier genes**

| **Gene** | **PEDAYA IQR** | **TCGA IQR** | **change in ped IQR relative to TCGA** | **PEDAYA median** | **TCGA median** | **change in ped median relative to TCGA** |
| --- | --- | --- | --- | --- | --- | --- |
| BCL6 | 2.18 | 1.53 | 0.43 | 4.3 | 4.42 | -0.03 |
| CCND3 | 2.14 | 1.1 | 0.94 | 6.04 | 5.54 | 0.09 |
| CDK9 | 1.84 | 0.73 | 1.53 | 5.7 | 5.28 | 0.08 |
| ETV1 | 4.1 | 2.22 | 0.85 | 1.82 | 2.23 | -0.18 |
| FGFR1 | 3.33 | 2.09 | 0.59 | 5.25 | 4.51 | 0.17 |
| FGFR2 | 4.2 | 2.3 | 0.82 | 1.34 | 3.47 | -0.62 |
| FGFR3 | 3.69 | 3.05 | 0.21 | 1.21 | 3.11 | -0.61 |
| GATA2 | 2.88 | 1.97 | 0.46 | 2.02 | 2.02 | 0 |
| HMOX1 | 3.11 | 1.8 | 0.73 | 3.74 | 4.69 | -0.2 |
| IGF2 | 6.28 | 2.92 | 1.15 | 2.85 | 3 | -0.05 |
| KDR | 3.17 | 1.72 | 0.84 | 2.03 | 2.5 | -0.19 |
| PARP2 | 1.21 | 0.98 | 0.24 | 4.69 | 4.04 | 0.16 |
| PDGFRA | 3.97 | 2.41 | 0.65 | 1.62 | 2.44 | -0.34 |
| PIK3CD | 2.85 | 1.56 | 0.83 | 3.23 | 2.43 | 0.33 |
| PTCH1 | 2.35 | 1.45 | 0.62 | 2.83 | 2.08 | 0.36 |
| TSC2 | 1.55 | 0.83 | 0.88 | 5.74 | 5.27 | 0.09 |
